# To isolate or not to isolate: The impact of changing behavior on COVID-19 transmission

**DOI:** 10.1101/2020.08.30.20184804

**Authors:** Folashade B. Agusto, Igor V. Erovenko, Alexander Fulk, Qays Abu-Saymeh, Daniel Romero-Alvarez, Joan Ponce, Suzanne Sindi, Omayra Ortega, Jarron M. Saint Onge, A. Townsend Peterson

**Affiliations:** University of Kansas, Lawrence, KS 66045 USA; University of Kansas Medical Center, Kansas City, KS 66160 USA; Purdue University, West Lafayette, IN 47907 USA; University of California Merced, Merced, CA 95343 USA; Sonoma State University, Rohnert Park, CA 94928; University of North Carolina at Greensboro, Greensboro, NC 27412 USA

**Keywords:** COVID-19, isolation and quarantine, game theory, human behavior, imitation dynamics, perception of risk

## Abstract

The COVID-19 pandemic has caused more than 25 million cases and 800 thousand deaths worldwide to date. Neither vaccines nor therapeutic drugs are currently available for this novel coronavirus. All measures to prevent the spread of COVID-19 are thus based on reducing contact between infected and susceptible individuals. Most of these measures such as quarantine and self-isolation require voluntary compliance by the population. However, humans may act in their (perceived) self-interest only. We construct a mathematical model of COVID-19 transmission with quarantine and hospitalization coupled with a dynamic game model of adaptive human behavior. Susceptible and infected individuals adopt various behavioral strategies based on perceived prevalence and burden of the disease and sensitivity to isolation measures, and they evolve their strategies using a social learning algorithm (imitation dynamics). This results in complex interplay between the epidemiological model, which affects success of different strategies, and the game-theoretic behavioral model, which in turn affects the spread of the disease. We found that the second wave of the pandemic, which has been observed in the US, can be attributed to rational behavior of susceptible individuals, and that multiple waves of the pandemic are possible if the rate of social learning of infected individuals is sufficiently high. To reduce the burden of the disease on the society, it is necessary to incentivize such altruistic behavior by infected individuals as voluntary self-isolation.

## 1 Introduction

COVID-19 is a respiratory disease caused by a recently discovered, novel coronavirus SARS-CoV-2. Since its discovery in Wuhan, China, in 2019, COVID-19 has led to over 25 million cases globally, over 800 thousand deaths, and 16 million recovered. Spreading globally, including to vulnerable countries with challenging healthcare infrastructures, the virus is now of international concern and has been deemed a pandemic by the World Health Organization (WHO).

According to COVID-19 data from Johns Hopkins University [31], the United States is currently the epicenter of the outbreak, with nearly 5 million confirmed cases and over 180 thousand reported deaths. Additionally, South America, India, and Africa are experiencing rising in cases and deaths from the virus. Brazil has over 3 million confirmed cases with over 120 thousand deaths; India has over 3 million confirmed cases with over 62 thousand deaths; and South Africa has over 600 thousand confirmed cases and 13 thousand deaths. These statistics point towards a grim realization that the world might be losing the battle to contain and control the pandemic.

COVID-19 is transmitted person-to-person *via* respiratory droplets and aerosols or by touching contaminated surfaces and objects containing the virus [5]; the virus can live for hours or days on contaminated surfaces and objects [10]. The incubation period for those exposed to COVID-19 varies from 2 to 12 days [11, 14]; onset of symptoms is often seen earlier in people with pre-existing health conditions and compromised immune systems. There is a wide range of symptoms observed in patients with COVID-19, including fever, shortness of breath, dry cough, headache, nausea, sore throat, chest pain, loss of taste or smell, diarrhea, and severe fatigue [14].

While the risk of severe complications and death from COVID-19 is higher among the older population and people with pre-existing conditions, younger adults and children remain at risk. In China, 90% of children were asymptomatic and only 5.9% had severe infections (compared to 20% among adults with the disease) [23]. In Italy, 10% of COVID-19 infected people in ICUs are 20–40 years old [16, 28]. Nonetheless, many young people are not taking the pandemic seriously [28]. In the United States, there have been numerous examples of young adults ignoring these warnings and underestimating the disease risk either to themselves or to older individuals around them. For instance, a group of young adults in Kentucky threw a Coronovirus Party [49] and other gathered in an over-crowded pool party without social distancing [21].

Since neither vaccines nor therapeutics are yet available for this virus, public health responses require social policies. Various regions have tried distinct responses including social distancing, school and event closings, and travel bans. Social distancing guidelines as suggested by the Centers for Disease Control and Prevention (CDC) and the World Health Organization states that individuals outside their homes should be six feet apart from all other people and to wear a face mask at all times. The guidelines further recommend that people frequently wash their hands for at least 20 seconds, even in their homes, as research has shown that soap kills the virus and reduces ones chance of getting infected [13]. Infected individuals and suspected cases are quarantined or advised to self-isolate. However, little is known about best management strategies for limiting further transmission and spread. Furthermore, the success of these preventive measures depend on voluntary compliance by the population, humans may act in their (perceived) self-interest only.

The objective of this study is to gain insight into the role of human behavior in modulating the spread and prevalence of COVID-19. We construct a mathematical model of COVID-19 transmission with quarantine and hospitalization, and we couple this model with a dynamic game model of adaptive human behavior. Susceptible individuals seek to protect themselves from the infection, and they consider supporting school and workplace closures. Infected individuals cannot protect themselves, but they may try to protect the rest of the population by electing to self-isolate from other people. Individuals adopt strategies based on the perceived prevalence and burden of the disease and on sensitivity to the social isolation measures. They may also imitate strategies of other individuals via a social learning process (imitation dynamics [29])if these individuals are more successful according to appropriately defined game payoff functions. This results in a complex interplay between the disease spread and human behavioral response, which affect each other in a feedback loop. We try to identify behavioral factors that reduce the scale of the pandemic, and propose possible measures to address these factors for the benefit of the entire society.

## 2 Results

We begin by analysing a baseline model ofCOVID-19transmission with quarantine and hospitalization (described in Section 4.1). We then analyze two models of dynamically adapting human behavior within the baseline model (described in Section 4.2): support for school and workplace closures by susceptible individuals to protect themselves from infection, and self-isolation by symptomatically infected individuals to protect others from infection. We analyze the effect of each type of behavior on the spread and prevalence ofCOVID-19 separately and jointly.

### 2.1 Baseline COVID-19 model

We construct a model of COVID-19 transmission with quarantine and hospitalization in Section 4.1. We follow the natural history of the infection [42, 53] and partition the population according to their disease status as susceptible (*S*(*t*)), exposed (*E*(*t*)), asymptomatically infected (*A*(*t*)), symptomatically infected (*I*(*t*)), quarantined (*Q*(*t*)), hospitalized (*H*(*t*)), and removed (*R*(*t*)) individuals. The flow diagram depicting the transition from one state to the other as the disease progresses through the population is shown in Figure 1, and the associated state variables and parameters are described in Tables 1.

**Figure 1:**
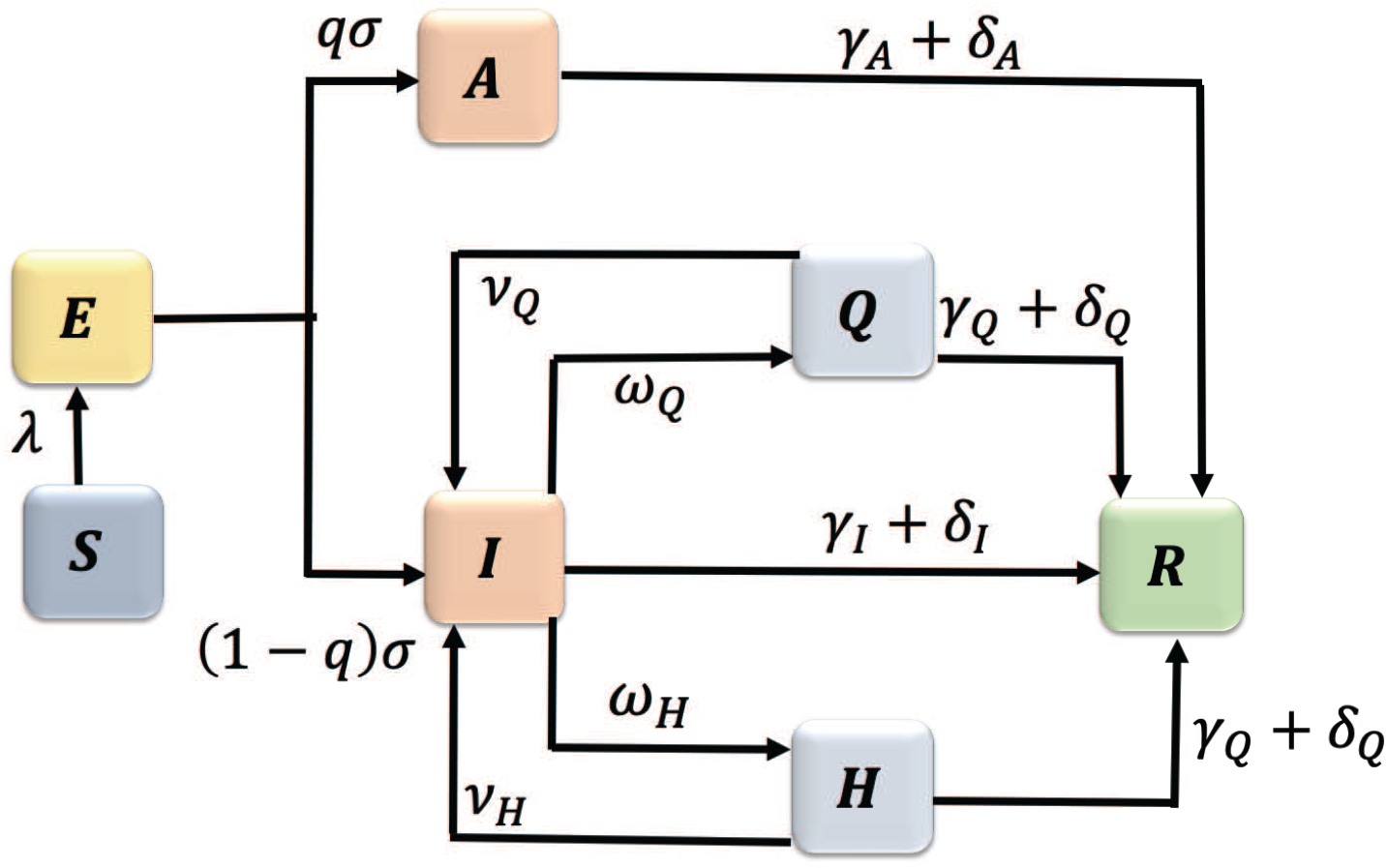
Flowdiagram of theCOVID-19 model(1).

**Table 1:** Description of the variables andparameters oftheCOVID-19 model(1).

| Variable | Description |
| --- | --- |
| $S(t)$ | Proportion of susceptible individuals |
| $E(t)$ | Proportion of exposed individuals |
| $A(t)$ | Proportion of asymptotically infected individuals |
| $I(t)$ | Proportion of symptomatically infected individuals |
| $Q(t)$ | Proportion of quarantined individuals |
| $H(t)$ | Proportion of hospitalized individuals |
| $R(t)$ | Proportion of removed individuals |
| Parameter | Description |
| $\beta$ | Infection rate |
| $\eta_A, \eta_Q, \eta_H$ | Modification parameters for asymptomatic, quarantined, and hospitalized infection rates |
| $q$ | Proportion of exposed developing asymptomatic infections |
| $\sigma$ | Disease progression rate from the exposed to infectious |
| $\gamma_I, \gamma_A, \gamma_Q, \gamma_H$ | Recovery rates of symptomatic, asymptomatic, quarantined, and hospitalized individuals |
| $\omega_Q, \omega_H$ | Quarantine and hospitalization rates |
| $\nu_Q$ | Quarantine violation rate |
| $\nu_H$ | Hospital discharge rate |
| $\delta_I, \delta_A, \delta_Q, \delta_H$ | Death rates of symptomatic, asymptomatic, quarantined, and hospitalized individuals |

The associated reproduction number[22,48] of thebaselineCOVID-19 model(1) with quarantine and hospitalization, denoted by 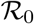, is given by

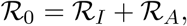

where

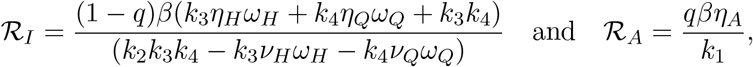

with *k*_1_ = *γ_A_* +*δ_A_*, *k*_2_ = *γ_I_* +*ω_Q_* +*ω_H_* +*δ_I_, k*_3_ = *ν_Q_* +*γ_Q_* +*δ_Q_*, and *k*_4_ = *ν_H_* +*γ_H_* +*δ_H_*. The quantity 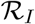 is the number of secondary infections produced by symptomatic individuals, while 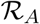 is the number of secondary infections generated by asymptomatic individuals. Together, the epidemiological quantity 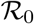, measures the average number of COVID-19 secondary infections produced when a single infected individual is introduced into a completely susceptible population [22,48]. Hence,COVID-19 can be effectively controlled in the population if the reproduction number 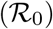 can be reduced to(and maintained at) a value less than unity(i.e., 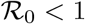).

We computed the numerical value of the reproduction number 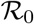 using the parameter values tabulated in Table 3. Some of the parameter values in Table 3 were fitted based ontheCOVID-19 data for Arizona from January 26 to July 6 [31] (see Figure 12), while others were obtained from the literature. Using these parameter estimates, we obtain 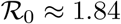 for the COVID-19 outbreak in Arizona.

#### 2.1.1 Impact of quarantine and hospitalization

Here, we investigate the impact of quarantine and hospitalization on the disease transmission. We vary the values of the quarantine rate *ω_Q_*, hospitalization rate *ω_H_*, quarantine violation rate *ν_Q_*, early discharge of symptomatic infectious individuals from hospitals rate *ν_H_*, and the infection rate *β* in pairs and examine the effect of these variations on the value of 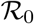.

Figure 2(a) shows that increasing quarantine and hospitalization rates reduces the value of 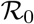, but the disease burden is still high because the values of 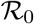 are greater than one. However, Figure 2(b) shows that the values of 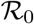 can be kept below 1 as long as the values of *β* do not exceed a certain threshold (*β* ≈ 0.22), and this outcome does not depend on the quarantine and hospitalization rates (see also Figure 13(a)). Using this lower level of the infection rate, we see in Figure 13(b) that 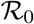 can be kept below 1 provided either the quarantine rate is above 0.4 or the hospitalization rate is above 0.2.

**Figure 2:**
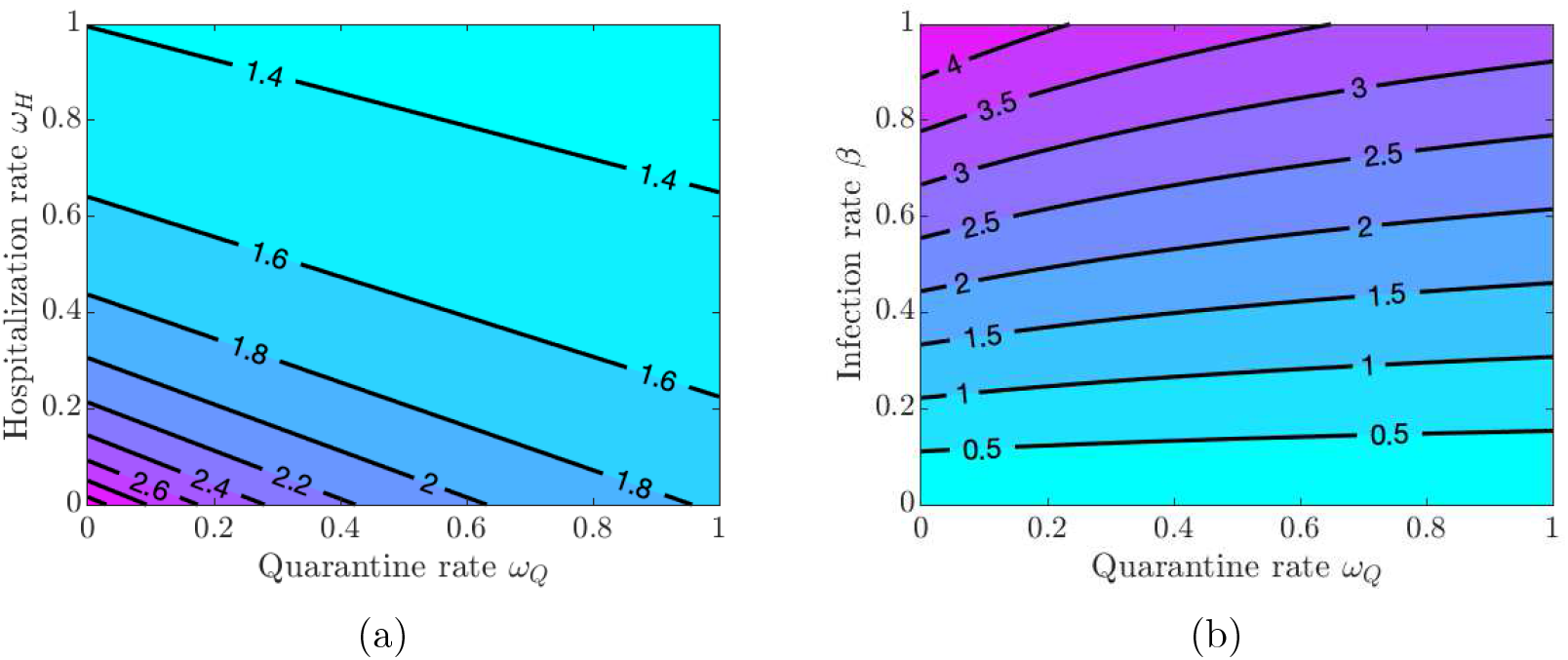
Contour plot of the COVID-19 reproduction number 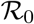 given in equation (1). (a) Varying quarantine rate *ω_Q_* and hospitalization rate *ω_H_*. (b) Varying quarantine rate *ω_Q_* and infection rate *β*.

If symptomatically infected individuals violate quarantine or are discharged from the hospitals into the community due to overwhelmed demand for hospitalizations or lack of resources, then the disease burden is high and containing the disease becomes challenging as values of 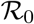 are greater than 1 for all values of *ν_Q_* and *ν_H_* (see Figure 3(a)). The situation is even worse if quarantine violation is varied along with poor hygiene and disregard for social distancing, which increases the infection rate *β*. Figures 3(b) and 14(a) show that 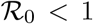 as long as the values of *β* do not exceed approximately the same threshold value *β* ≈ 0.22 as in the case of varying quarantine and hospitalization rates. Figure 14(b) shows that the values of 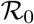 are below 1 provided the quarantine violation rate *ν_Q_* is below 0.7 or the hospital discharge rate *ν_H_* is below 0.4. Moreover, 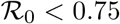 if both *ν_Q_* and *ν_H_* are below 0.2.

**Figure 3:**
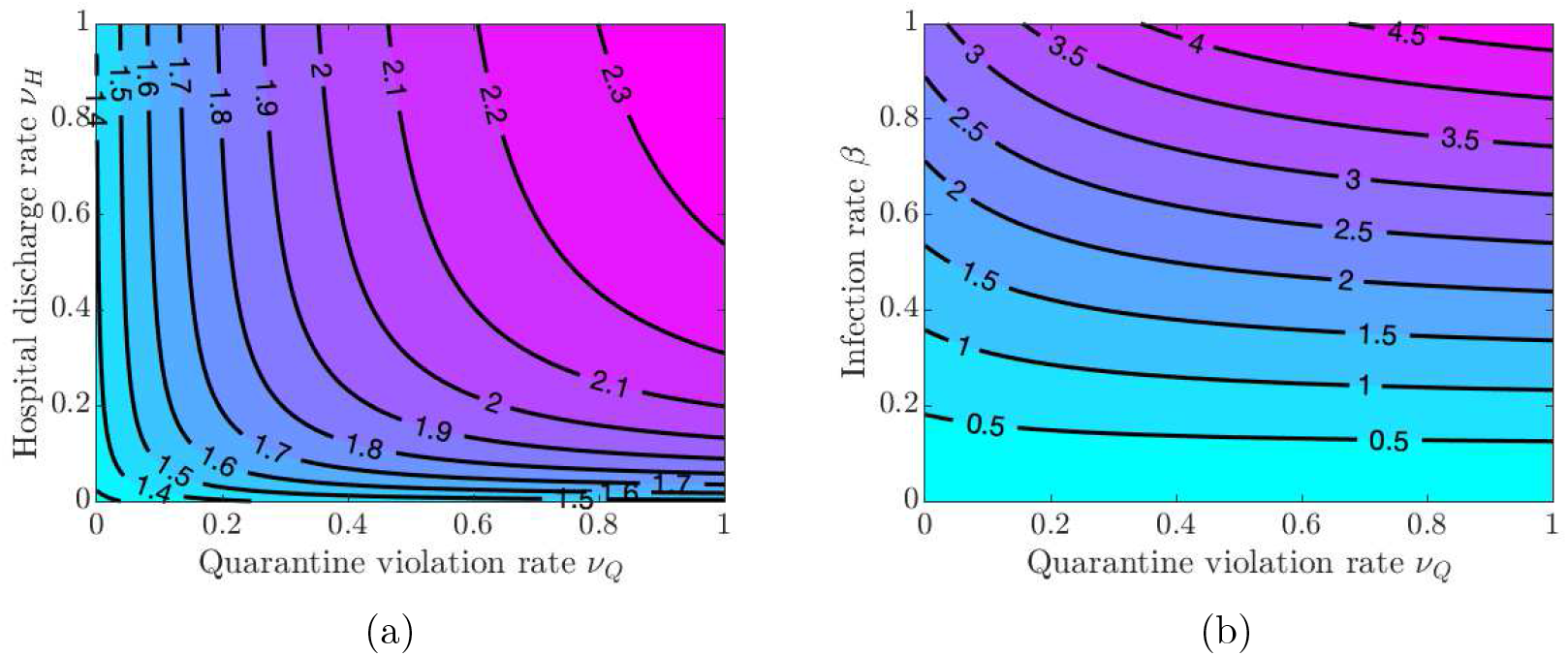
Contour plot of the COVID-19 reproduction number 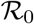 given in equation (1). (a) Varying quarantine violation rate *ν_Q_* and hospital discharge rate *ν_H_*. (b)Varying infection rate *β* and quarantine violation rate *ν_Q_*.

The results in Figures 2 and 3 show the importance of keeping the infection rate *β* low in order to reduce the disease burden. This can be achieved by maintaining proper hygiene(washing hands as recommended), social distancing, and using facial masks.

#### 2.1.2 Role of quarantined and hospitalized individuals

In this section, we investigate the impact of quarantine and hospitalization on the proportion of infected individuals that exhibit symptoms of COVID-19. These individuals span three compartments: *I*, *Q*, and *H*. Figure 4(a) shows the effect of doubling the quarantine (*ω_Q_*) and hospitalization (*ω_H_*) rates. The overall number of infections is reduced, and the epidemic curve is flattened, while the peak of the infection is shifted to later in time. On the other hand, doubling the quarantine violation (*ν_Q_*)and hospital discharge (*ν_H_*) rates results in a higher infection peak that occurs sooner; see Figure 4(b). These simulations further suggest, as expected, that a larger COVID-19 burden would be recorded if more people violate the quarantine rules, while increasing the quarantine rate lowers the disease burden in the community.

**Figure 4:**
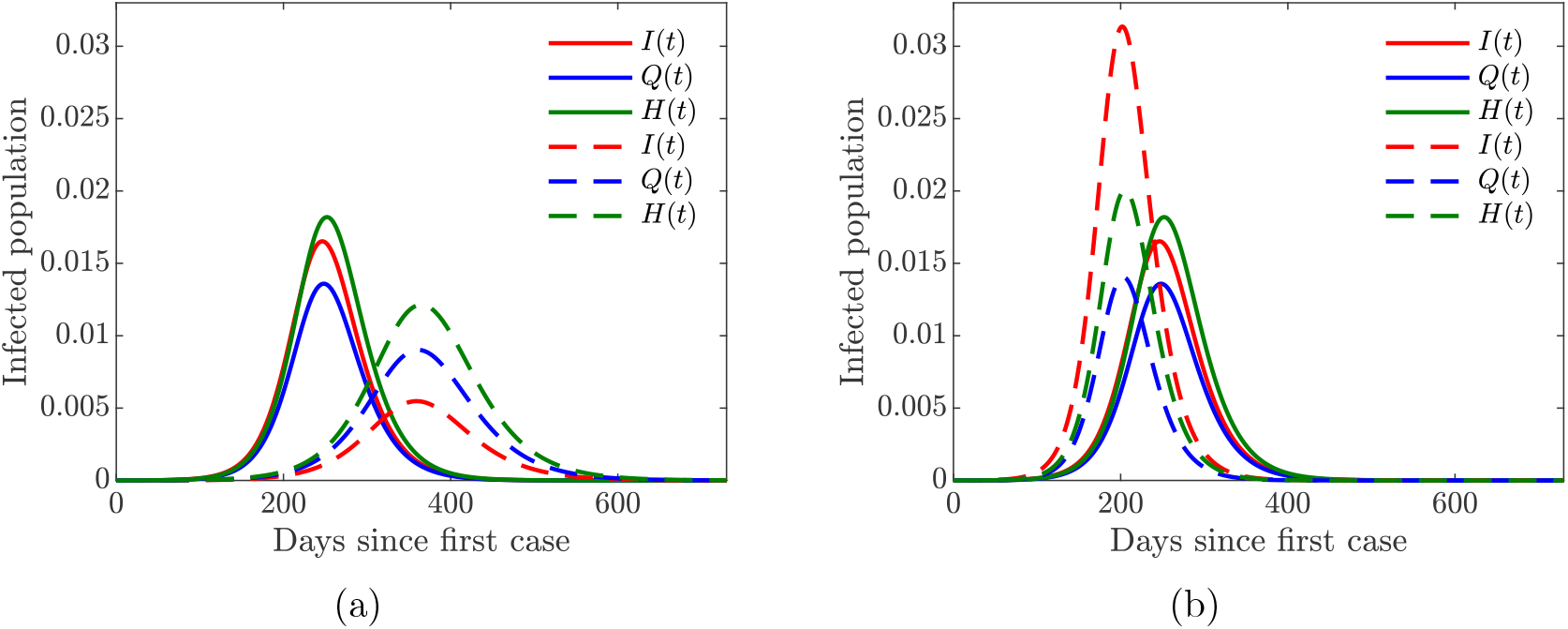
Simulation of the baseline COVID-19 model (1) for the proportions of symptomatically infected (*I*), quarantined (*Q*), and hospitalized (*H*) individuals. Solid lines correspond to base values of the model parameters from Table 3. (a) Dashed lines correspond to double quarantine (*ω_Q_*) and hospitalization (*ω_H_*) rates (b) Dashed lines correspond to double quarantine violation (*ν_Q_*) and hospital discharge (*ν_H_*) rates.

In summary, the simulations of the COVID-19 model (1) with static human behavior show that:

i. Increased quarantine violation and hospital discharge rates of those still infectious due to overwhelmed hospital resources increases the disease burden leading to an early epidemic peak.
ii. Increasing quarantine and hospitalization rates decreases the disease burden and reduces the epidemic peak. Moreover, these measures postpone the peak of the infection, thus giving more time to prepare for the coming spike of the disease.

### 2.2 The COVID-19 model with dynamic human behavior

Perceived risk of infection drives human behavior and decisions during an epidemic. These behaviors and decisions are derived from evaluating alternative decisions and weighing related cost-benefit [44]. In this section, we analyze the effects of dynamically changing human behavior by susceptible and symptomatically infected individuals within the baseline COVID-19 model (1); the extended model is given by equations (15). Unlike previous analyzes which focused on how susceptible individuals change their behavior related to the use and acceptance of public health protective and preventive control measures [2, 3, 41, 43, 54], we also consider change in behavior and decision making of the downstream symptomatically infected population. The state variables and parameters associated with the behavioral model are summarized in Table 2.

#### 2.2.1 Susceptible support for closure

We begin by analyzing the effect of the susceptible individuals support or opposition of school and workplace closures. To isolate the effect of susceptible individual behavior, we assume that *κ_I_* = 0 and *x_I_*(0) = 0, that is, the symptomatically infected individual behavior is suppressed. Our modeling approach to the susceptible individual behavior is derived from [41], and is described in section 4.2.1. Susceptible individuals seek to avoid getting infected, and they weigh perceived risk of infection versus the possible socio-economic losses due to the partial economy shutdown; the socio-economic losses accumulate over time. We assumed that the decision to enact appropriate closures stays in effect if and only if a certain minimum time has passed since the start of the pandemic and at least half of the susceptible population supports closures.

In all simulations involving susceptible individual support for closure, we assume that the effectiveness of closures is *C*_0_ = 0.6 and the initial time the closure decision may be enacted is *t*_close_ = 30 days. Figure 5(a) shows the effect of dynamically changing susceptible individual behavior on the progression of the epidemic with different starting conditions, which capture the initial predisposition of the population towards such drastic measures as school and workplace closures.

**Figure 5:**
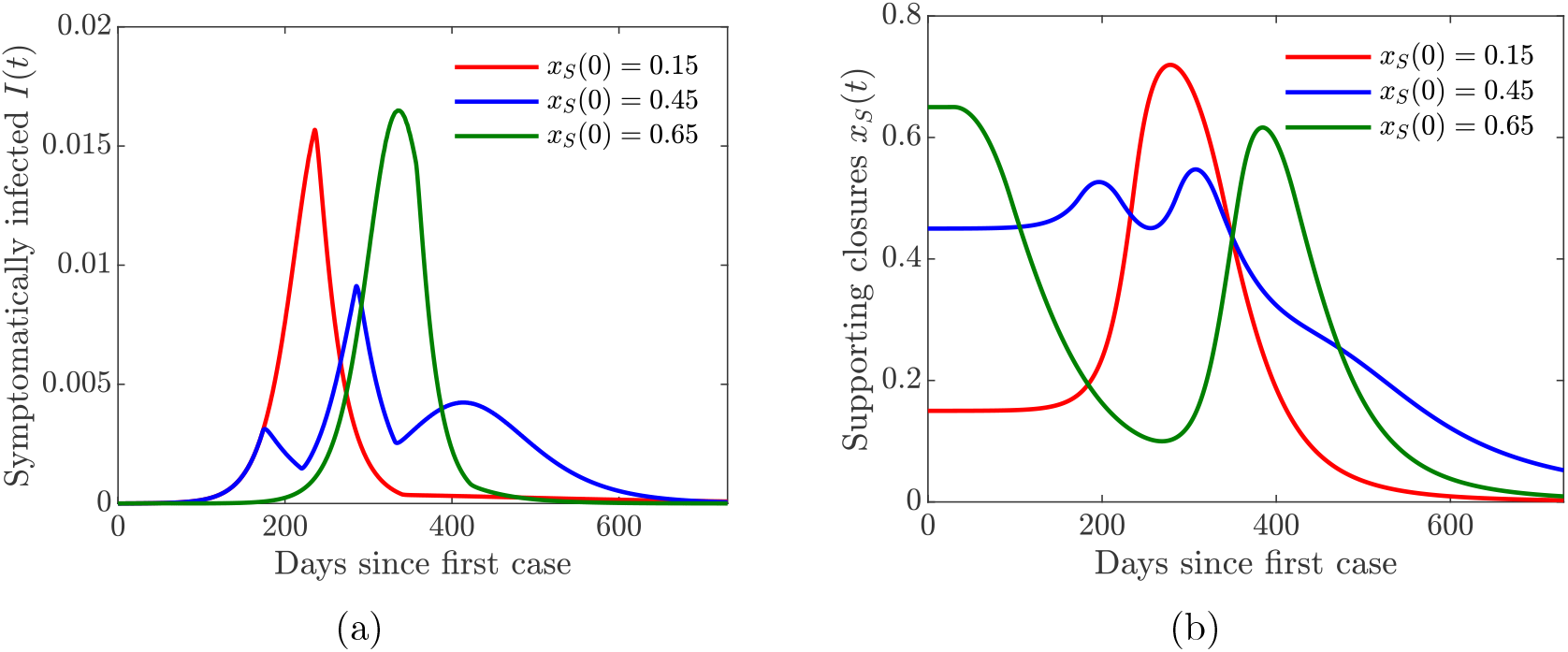
Simulations of the COVID-19 model with dynamic human behavior (15) with various initial proportions *x_S_*(0) of the susceptibles in support of lock-down. The social learning rate of susceptible individuals is *κ_S_* =1. (a) The progression of the proportion of symptomatically infected individuals *I*(*t*). (b) The progression of the proportion of the susceptible population in support of the closure or lock-down measures. The measures are enacted as long as *t* ≥ *t*_close_ and *x_S_* (*t*) ≥ 0.5.

When the population is initially skeptical about the closures (*x_S_* (0)=0.15), then it takes a while to build sufficient support for the measure to be enacted (Figure 5(b), red line). As a result, the closures take place too late, and the pandemic reaches its peak early on (Figure 5(a), red line). On the other hand, when the population is initially overenthusiastic about the closures (*x_S_* (0) =0.65), the measure is enacted too early (Figure 5(b), green line). However, the accumulating socio-economic losses due to the lock-down start to wear people down, and the majority of the population begins to oppose the lock-down. This results in a sharp peak of the cases (Figure 5(a), green line), which is simply delayed in time. The rise in the prevalence of infection forces individuals to revert to the lock-down measures, but this switch in behavior comes too late to prevent a spike in infections.

The lowest infection peaks are achieved when the proportion of susceptible individuals initially supporting the closures is neither too low or too high but “just right” (*x_S_* (0) =0.45). The lock-down is enacted as soon as the number of cases begins to increase (Figures 5(a) and 5(b), blue lines). The initial epidemic is stifled, and the closure support drops below the threshold, which results in (partial) re-openings. However, the number of infected individuals is still relatively high, and a second bigger wave of infections occurs. The second wave forces another shutdown, which persists for a shorter period of time compared to the first one. This scenario is similar to what has been happening in the US, and it shows that a second wave of COVID-19 may result from rational human behavior due to the burden of accumulating socio-economic losses. This observation matches the results in [41], and it shows that our extended model with quarantine and hospitalization still captures the basic features of a simpler model.

For simplicity, we used only one value of the susceptible individual social learning rate parameter (*κ_S_* = 1) here. We investigate the effects of varying this parameter when we analyze a coupled model of susceptible and infected individual behavior. In particular, faster social learning rates may result in multiple waves of infection.

#### 2.2.2 Symptomatically infected self-isolation

We now analyze the effect of voluntary decisions to self-isolate by symptomatically infected individuals. We assume that *κ_S_* =0 and *x_S_* (0)=0 so that susceptible individual support for closure behavior is suppressed. Our modeling approach to symptomatically infected individual behavior is described in Section 4.2.2. Unlike susceptible individuals, who seek to protect themselves from the infection, infected individuals cannot protect themselves—they are already infected. However, conscientious individuals may wish to protect the rest of the population from getting infected; these individuals weigh the perceived burden of infecting others versus the inconvenience and cost of self-isolation.

Figure 6(a) shows the impact of dynamically changing infected individual behavior to self-isolate or not self-isolate on the progression of the epidemic. At the onset of the epidemic, when the number of cases and fatalities is relatively small, infected individuals would tend not to engage in voluntary self-isolation (Figure 6(b)). As the number of infections—and hence disease-induced deaths—grows, the burden on the susceptible population becomes larger, and the infected individuals are more willing to self-isolate to protect others. The initial predisposition of the population to the altruistic act of self-isolation determines the peak of the epidemic and its timing (Figure 6(a)). The more individuals are willing to self-isolate, the lower the peak and the later it occurs.

**Figure 6:**
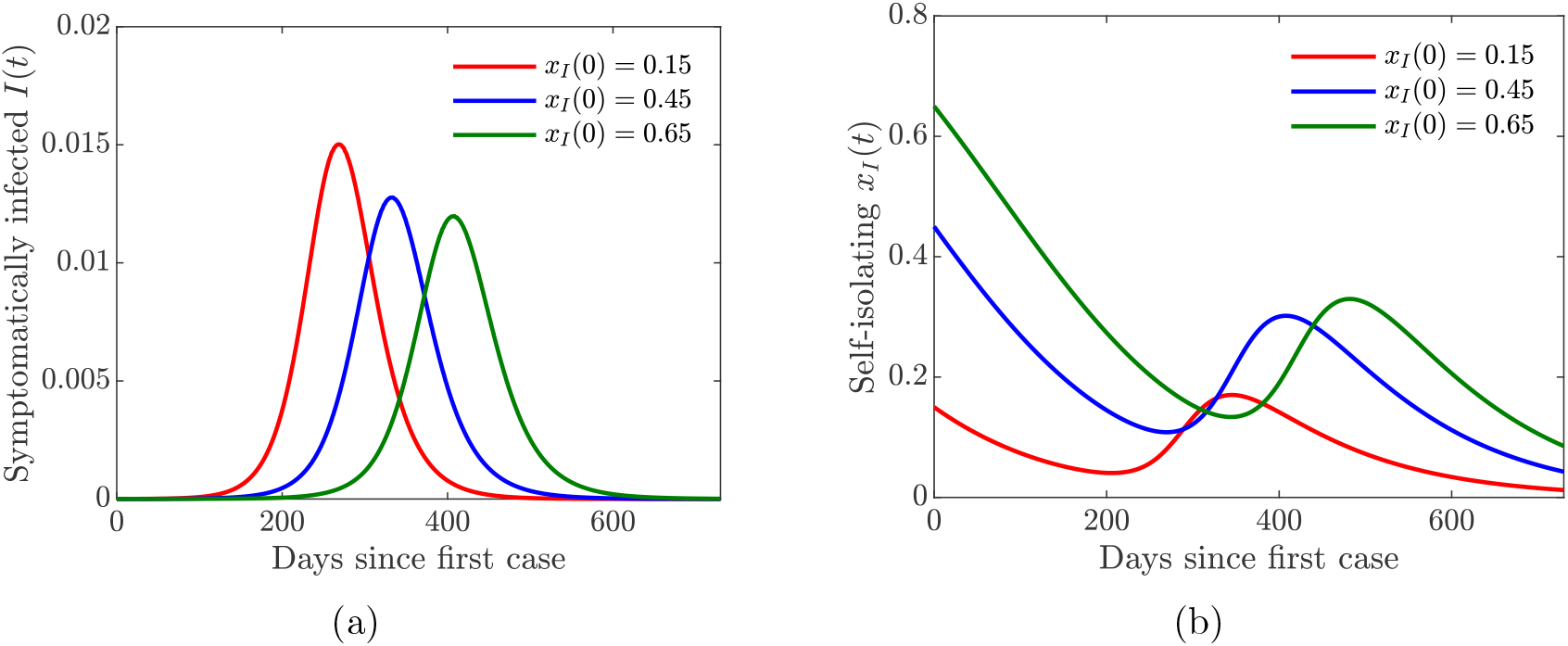
Simulations of the COVID-19 model with dynamic human behavior (15) with various initial proportions *x_I_* (0) of symptomatically infected individuals willing to self-isolate. The social learning rate of infected individuals is *κ_I_* =100, and the sensitivity to self-isolation is *ε_I_* =0.00008. (a) The progression of the proportion of symptomatically infected individuals *I*(*t*). (b) The progression of the proportion of symptomatically infected population willing to self-isolate.

We considered one set of fixed values of the symptomatically infected individual social learning rate parameter *κ_I_* and the sensitivity to self-isolation parameter *ε_I_*. We investigate the effects of varying these parameters in a full behavioral model. In particular, lowering the sensitivity to self-isolation results in bigger and more sustained support of self-isolation.

#### 2.2.3 Human behavior coupled with quarantine and hospitalization

In this section, we consider the full behavioral model, where both susceptible and symptomatically infected individuals adjust their behavior in response to the epidemic. We initialize the model simulations with only 15% of the susceptible population supporting closure and 15% of the symptomatic population willing to self-isolate, which correspond to the worst-case scenarios considered in Figures 5 and Figure 6.

Figure 7 shows the results of the simulation with varying quarantine (*ω_Q_*), hospitalization (*ω_H_*), quarantine violation (*ν_Q_*), and hospital discharge (*ν_H_*) rates. The peak of the epidemic is lower and shifted to the right in time with higher quarantine and hospitalization rates (Figure 7(a)), while an opposite effect is achieved with higher quarantine violation and hospital discharge rates (Figure 7(c)). The population behavioral response is informed by the severity of the epidemic: higher prevalence of the disease results in larger proportions of individuals supporting closure or willing to self-isolate (Figures 7(b) and 7(d)).

**Figure 7:**
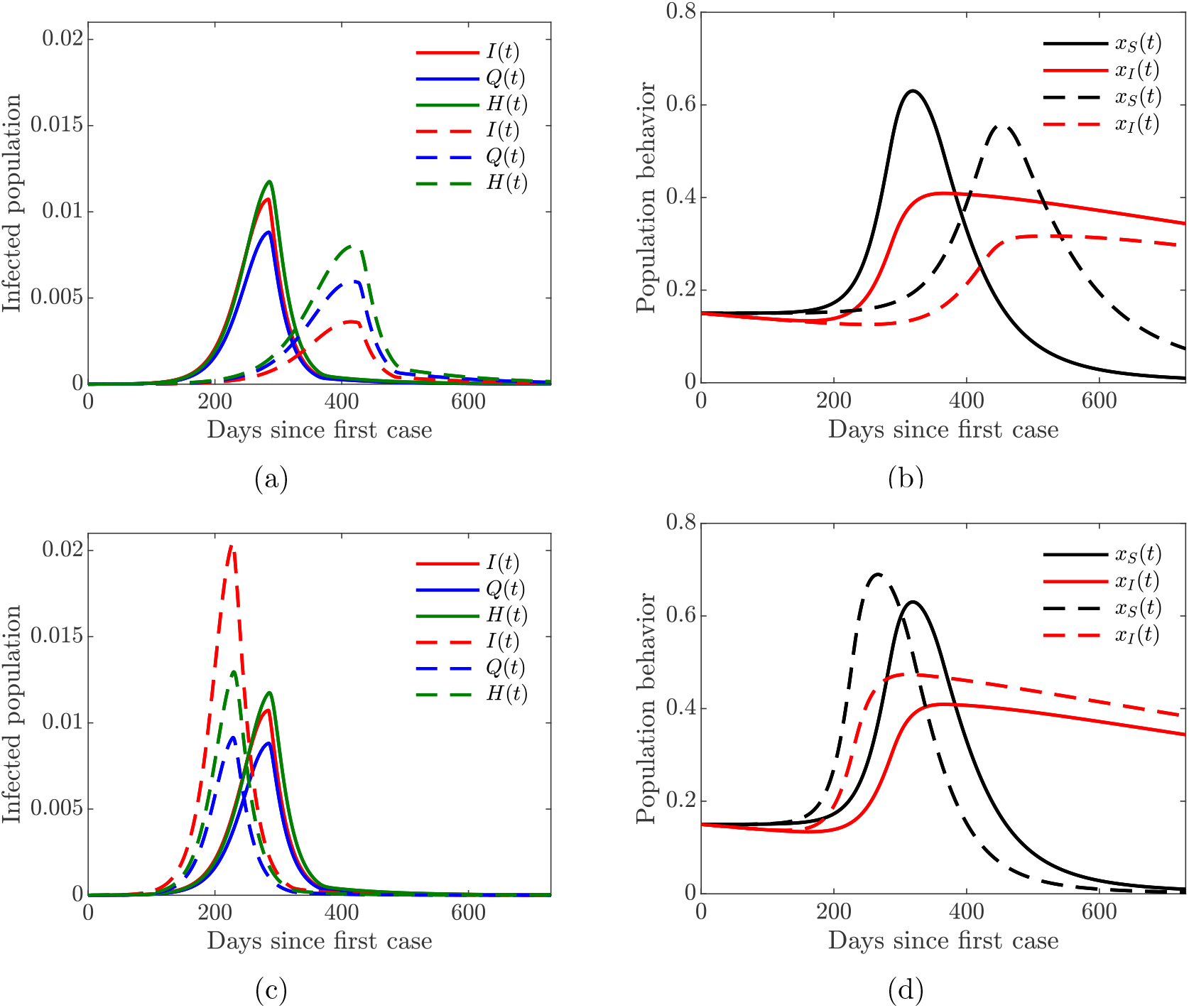
Simulations oftheCOVID-19 model with dynamic human behavior (15) for the proportions of all symptomatic infections and behavioral response with low sensitivity to self-isolation *ε_I_* =0.00001. The social learning rates are *κ_S_* = 1 and *κ_I_* =100, and *x_S_* (0)= *x_I_* (0)=0.15. Solid lines correspond to the values of the baseline model parameters given in Table 3. (a)–(b) Dashed lines correspond to double quarantine (*ω_Q_*) and hospitalization (*ω_H_*) rates (c)–(d) Dashed lines correspond to double quarantine violation (*ν_Q_*) and hospital discharge (*ν_H_*) rates.

Figure 7 illustrates the importance of discouraging disease-magnifying behavior such as violating and breaking quarantine laws. Moreover, lower sensitivity to self-isolation (*ε_I_* =0.00001 in Figure 7 compared to *ε_I_* =0.00008 in Figure 15) allows the self-isolating behavior to persist for a longer period of time (compare Figure 7(b) with 15(b) and Figure 7(d) with 15(d)) thus effectively reducing the burden of the infection on the susceptible part of the population (compare Figure 7(a) with 15(a) and Figure 7(c) with 15(c)). It is therefore important to encourage and incentivize such exemplary behavior by infected individuals.

### 2.3 Multiple waves of infections

In this section, we demonstrate the possibility of multiple waves of infection as a consequence of modifying the rates of behavioral response to the emerging epidemic conditions. The rates of behavioral response are controlled by the social learning rate parameters *κ_S_* and *κ_I_* for susceptible and symptomatically infected individuals, respectively, in the imitation dynamics model. Higher values of these parameters mean individuals imitate the behavior of other individuals, who are more successful according to the dynamic game payoffs, more eagerly. This effects quicker response to the evolving conditions, which may result in multiple oscillations of both the behavioral response and infections curves.

In general, we assumed that *κ_S_ <κ_I_* because supporting school and workplace closures usually carries bigger concessions than self-isolation. For example, individuals who can continue working remotely are more likely to support such measures, while individuals who will lose their jobs while part of the economy is shutdown are less likely to support measures that may result in loss or reduction of their income. Therefore, susceptible individuals may have different sensitivity to the socio-economic losses, and that is why we assumed that the social learning rate *κ_S_* for closure support behavior is lower than that for self-isolating behavior (*κ_I_*).

Figure 8 shows that increasing the support closure behavior social learning rate *κ_S_* produces oscillations in the behavioral response and hence in the prevalence of the disease. For higher values (*κ_S_* =30, see Figure 8(c)), we observe two waves of infections of similar magnitude. On the other hand, simultaneous increase in the self-isolation behavior social learning rate (*κ_I_* = 650) coupled with low sensitivity to self-isolation (*ε_I_* =0.00001) allows the population to overcome a second large wave of the pandemic by responding quickly and decidedly to the first big wave (see dashed lines in all panes of Figure 8). Still, increasing the self-isolation social learning rate parameter does not prevent a second large wave of the pandemic if the population sensitivity to self-isolation is higher (*ε_I_* =0.00008), see Figure 16.

**Figure 8:**
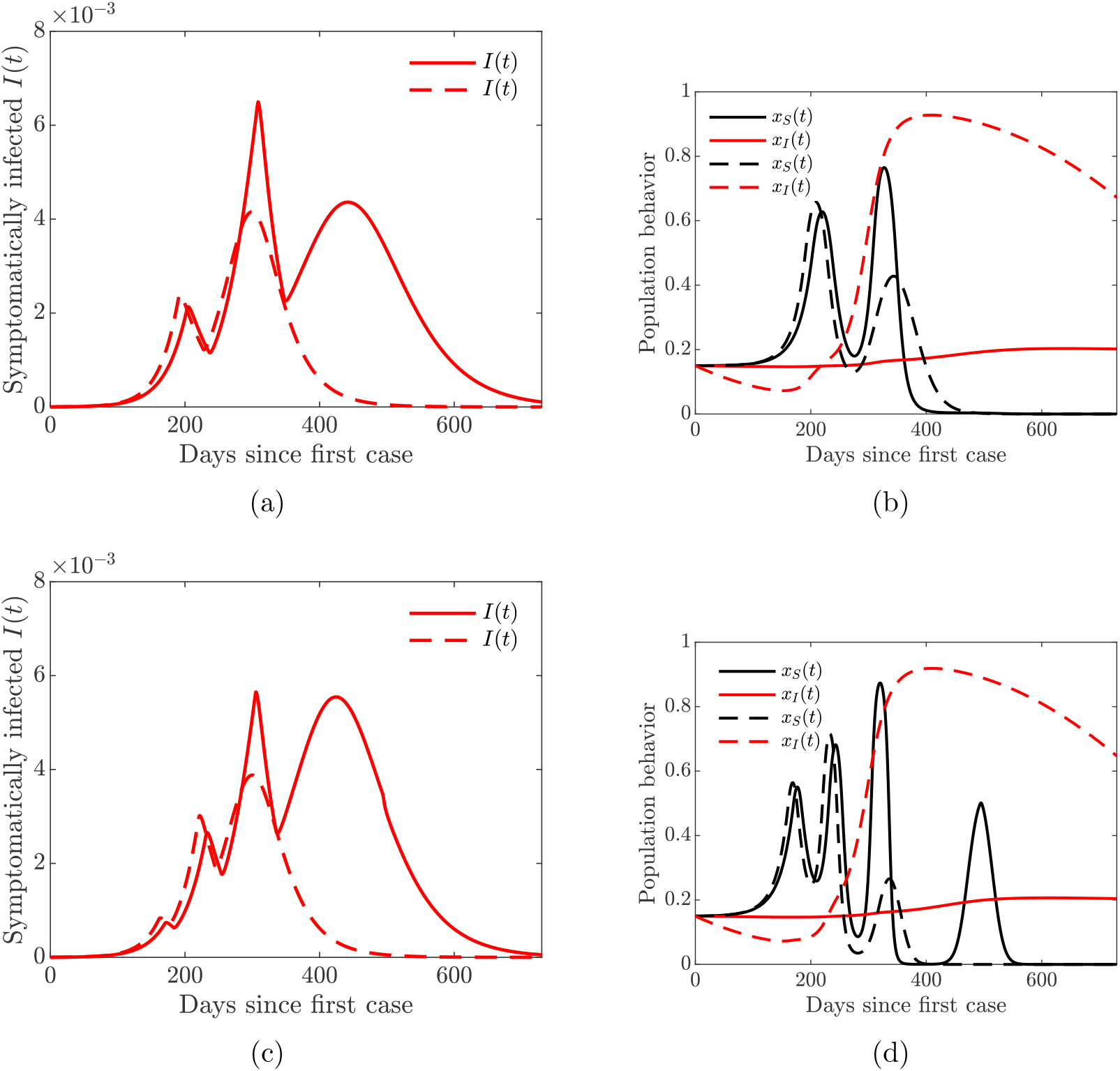
Simulations oftheCOVID-19model with dynamic human behavior (15) showing multiple waves of epidemic while varying susceptible (*κ_S_*) and symptomatic (*κ_I_*)individual social learning rates with low sensitivity to self-isolation *ε_I_* =0.00001. Solid lines correspond to *κ_I_* = 20, dashed lines correspond to *κ_I_* =650. (a) Proportion of symptomatic infections *I*(*t*) with one big and two smaller waves (solid lines), *κ_S_* = 10. (b) Proportion of susceptible (*x_S_*) and symptomatic (*x_I_*) individuals adopting positive behavior, *κ_S_* = 10. (c) Proportion of symptomatic infections *I*(*t*) with two big and one small wave (solid lines), *κ_S_* = 30. (d) Proportion of susceptible (*x_S_*) and symptomatic (*x_I_*) individuals adopting positive behavior, *κ_S_* =30.

Multiple waves of infection of similar magnitude may occur if the closure support social learning rate is low (*κ_S_* = 5) while the self-isolation social learning rate is high (*κ_I_* =1350) and sensitivity to self-isolation is low (*ε_I_* =0.00001); see Figure 9. This may seem counter-intuitive because higher willingness to self-isolate should ideally result in quick suppression of a spike in disease. At the same time, with high sensitivity to the epidemiological situation, individuals switch back to non-compliance as soon as the situation improves but well before the disease prevalence is reduced to negligible numbers. This, in turn, results in a new spike of infections. We note that this phenomenon is amplified by the presence of quarantine violation in our model because quarantine violation often results in outbreaks [4]. When the quarantine violation rate *ν_Q_* is set to zero, we no longer observe multiple epidemic waves of such magnitude.

**Figure 9:**
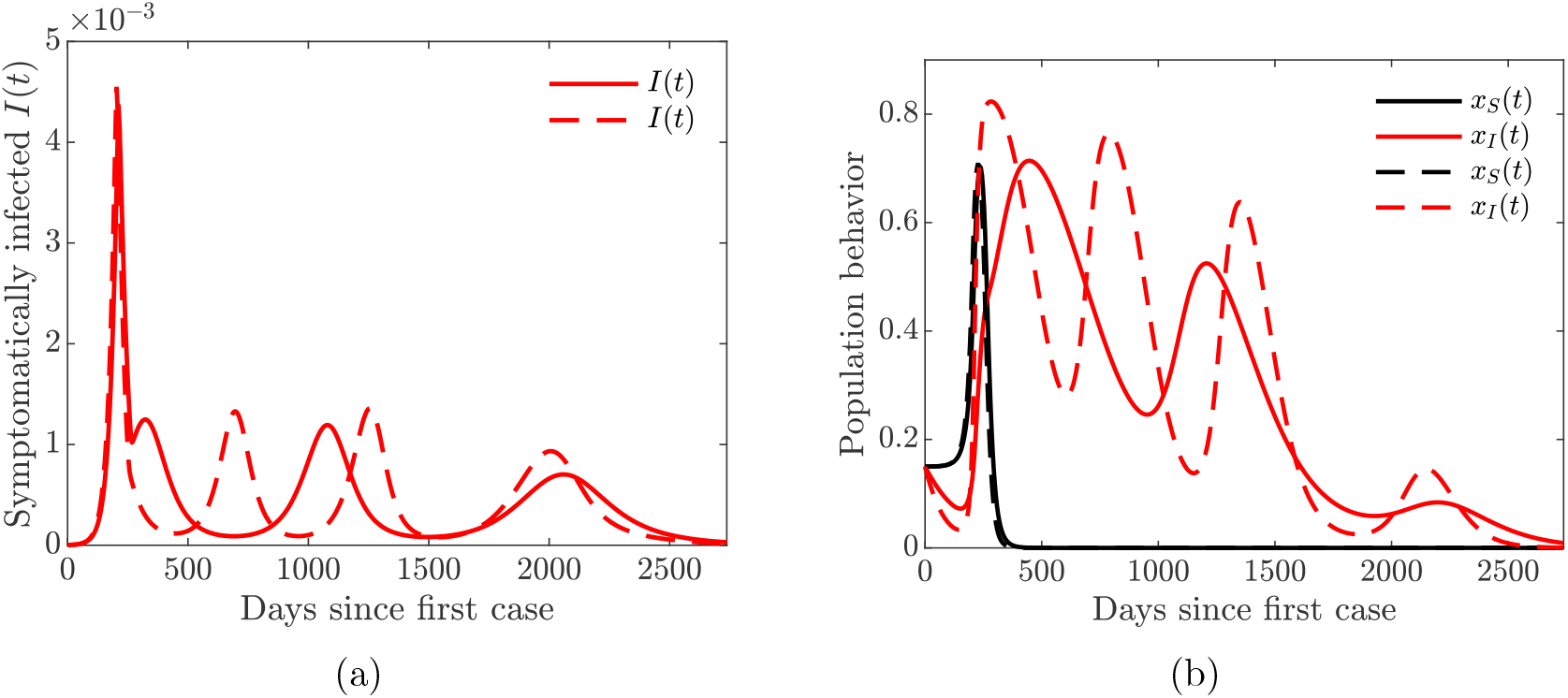
Simulations of theCOVID-19 model with dynamic human behavior (15) showing epidemic oscillations with high self-isolation social learning rate. Solid lines correspond to *κ_I_* = 650, dashed lines correspond to *κ_I_* = 1350; fixed values *κ_S_* = 5 and *ε_I_* = 0.00001. (a) Oscillating proportion of symptomatic infections *I*(*t*). (b) Proportion of susceptible (*x_S_*) and symptomatic (*x_I_*) individuals adopting positive behavior.

Figures 8 and 9 show the possibility of multiple epidemic waves or an epidemic with several oscillations. We have seen that the persistence of these waves is due to the high rate of social learning behavior of the susceptible or symptomatically infected individuals in the community or the violation of the quarantine rules. We will now explore in more detail the impact of increased quarantine and quarantine violation rates on the multiple epidemic waves. We will couple this with varying hospitalization and hospital discharge rates.

Figure 10 shows that increasing the quarantine and hospitalization rates prevents future waves of infection. This is achieved by dampening multiple oscillations in the behavior of symptomatically infected individuals and prolonged support for lock-down measures.

**Figure 10:**
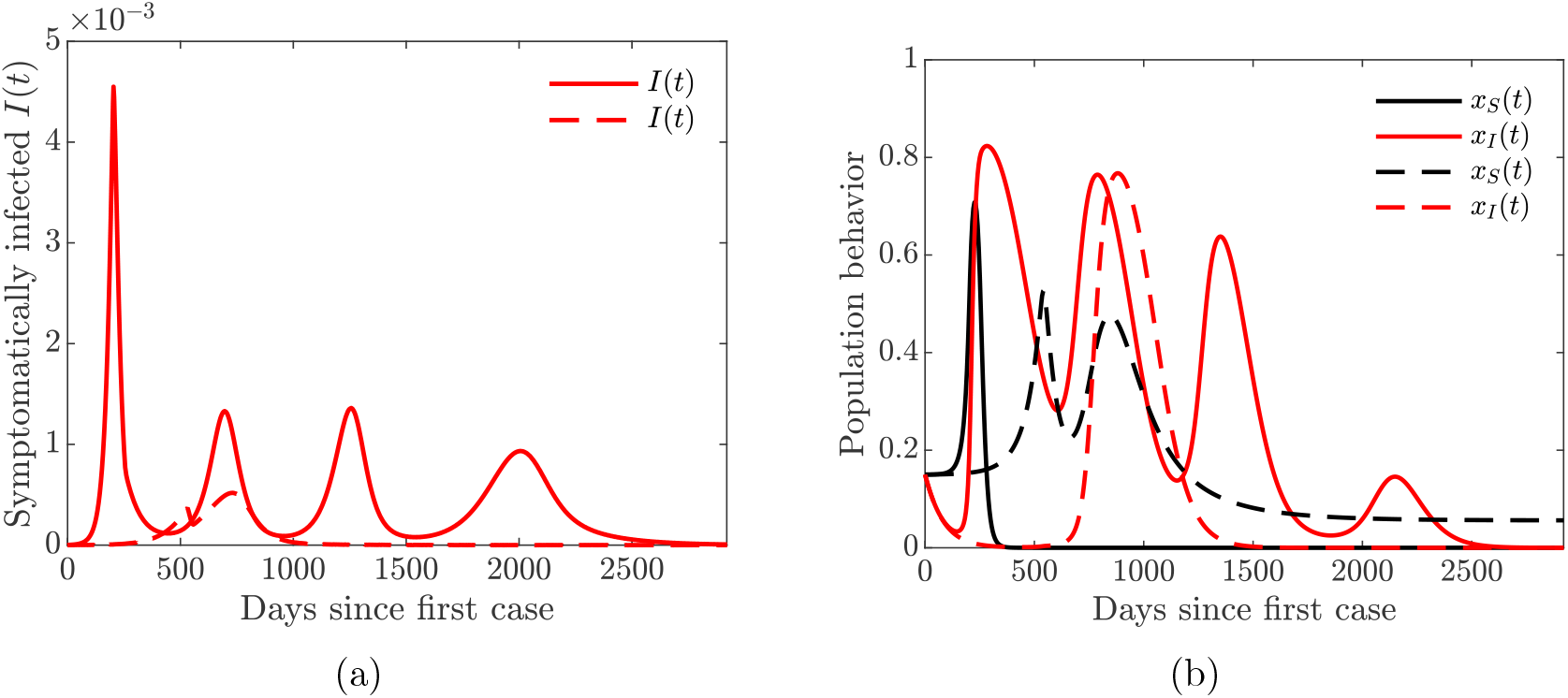
Simulations of theCOVID-19 model with dynamic human behavior (15) showing the damping effect of increased quarantine (*ω_Q_*) and hospitalization (*ω_H_*)rates. Solid lines correspond to base values of *ω_Q_* and *ω_H_*, dashed lines correspond to a 5-fold increase in these values; fixed values *κ_S_* = 5, *κ_I_* = 1350, and *ε_I_* =0.00001. (a) Proportion of symptomatic infections *I*(*t*). (b) Proportion of susceptible (*x_S_*)and symptomatic (*x_I_*)individuals adopting positive behavior.

Lastly, we investigate the impact of increased quarantine violation and hospital discharge rates on multiple waves of infection. We see from Figure 11 that increasing quarantine violation and hospital discharge rates produces multiple epidemic peaks of larger magnitude. Higher initial prevalence of the disease (Figure 11(a) dashed line) causes multiple oscillations in self-isolating behavior (Figure 11(b)) and hence future waves of infection.

**Figure 11:**
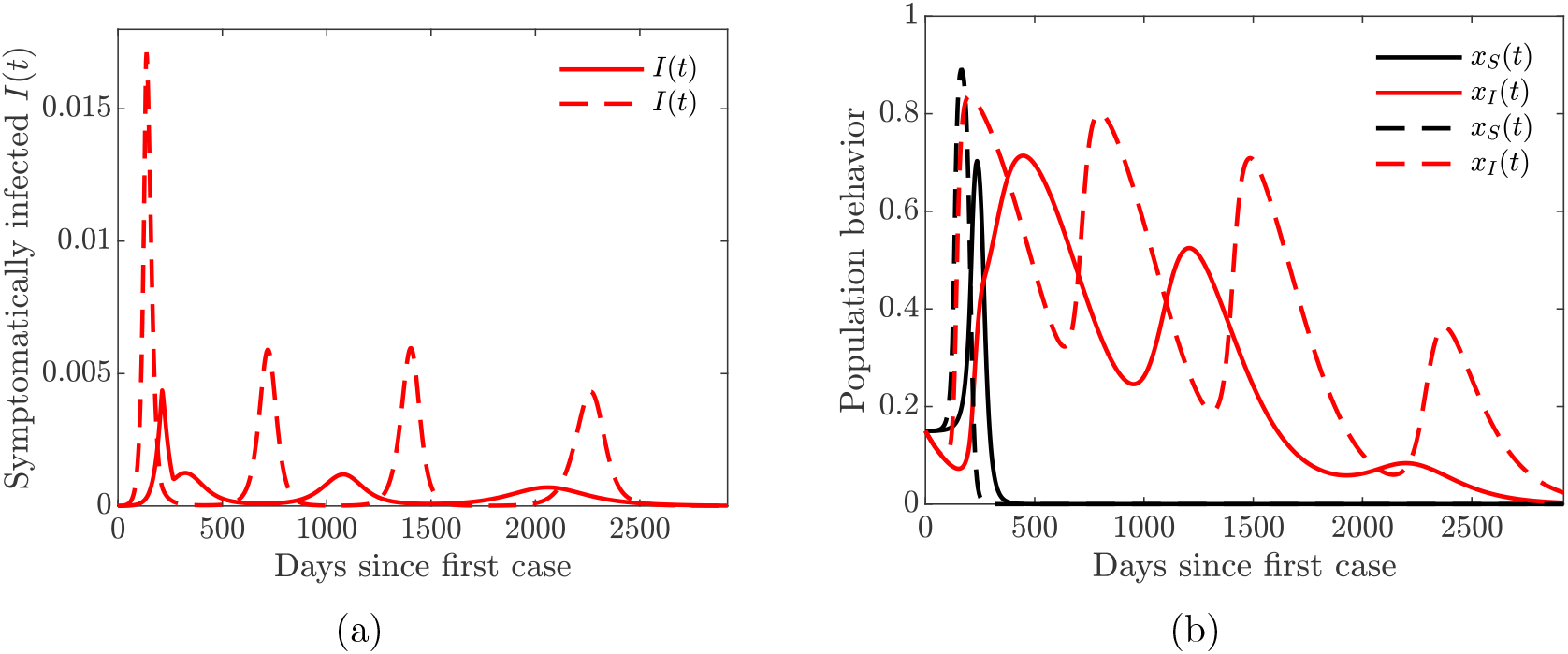
Simulations of theCOVID-19 model with dynamic human behavior (15) showing the devastating effect of increased quarantine violation (*ν_Q_*) and hospital discharge (*ν_H_*) rates. Solid lines correspond to base values of *ν_Q_* and *ν_H_*, dashed lines correspond to an 8-fold increase in these values; fixed values *κ_S_* = 5, *κ_I_* = 650, and *ε_I_* =0.00001. (a) Proportion of symptomatic infections *I*(*t*). (b) Proportion of susceptible (*x_S_*) and symptomatic (*x_I_*) individuals adopting positive behavior.

The take home-message from the results presented in Figures 10 and 11 is that increased hospitalization and quarantine rates can help diminish future infection waves and could even lead to the disappearance of a second large wave. However, frequent quarantine violation and early hospital discharge of those still infectious may lead to persistent prevalence of the disease with regular spikes in the number of cases.

In summary, the simulations oftheCOVID-19model with dynamic human behavior (15)show that:

i. Symptomatic individuals learning and mimicking self-isolating behavior reduces the disease burden in the population but can lead to multiple epidemic waves if fewer susceptible individuals mimic and learn closure support behavior.
ii. Quarantine violation and hospital discharge of symptomatic individuals amplifies the peaks of the infection waves and can lead to infection waves that persist in the community.
iii. Increasing quarantine and hospitalization rates can prevent multiple waves of infection.
iv. It is important to incentivize the cost and burden of self-isolation to encourage more symptomatic individuals to self-isolate because high sensitivity to self-isolation is not beneficial to the community as a whole.

## 3 Discussion and Conclusions

### 3.1 Discussion

We constructed a novel compartmental model of COVID-19 transmission, which includes compartments for quarantined and hospitalized individuals; see Figure 1 and equations (1). We coupled this model with a game-theoretic model of dynamically changing human behavior in equations (15). The susceptible individuals choose to either support school and workplace closures or not, and their strategic choices are driven by the perceived risk of getting infected versus the sensitivity of possible socio-economic losses due to the (partial) lock-down. The symptomatically infected individuals consider protecting the rest of the population by self-isolating from society; they base their decisions on the perceived burden of the disease versus the burden of social isolation.

We also investigated the effects of quarantine violation due to social non-compliance and early hospital discharge due to shortage of resources. Increasing the rates of quarantine violation and hospital discharge results in a higher peak of the pandemic, which occurs earlier (Figure 4) and hence could be more devastating. At the height of the outbreak in Michigan and New York, hospitals were discharging early the not-too-critically ill either to nursing homes or simply letting them go home because hospital facilities were overwhelmed[36,45]. This prompted legislation in Michigan to protect the seniors and vulnerable members of the community and prevent nursing homes from admitting patients with COVID-19 [37]. In other places like Arizona, some nursing homes are actually taking COVID-19patients with mild symptoms[20].

To reduce the disease burden in the community, it is important to keep the infection rate *β* low (approximately 0.22). This can be achieved by maintaining proper hygiene (frequently washing hands for 20 seconds), social distancing, and wearing facial masks. Unfortunately, the use of facial masks has become a polarizing topic in the United States, resulting in shaming, and violence [6, 33, 35, 46]. Nevertheless, the science behind the use of facial masks shows that the use of surgical masks prevent the dispersal and transmission of COVID-19 droplets and aerosols [8, 18, 34], and hence using facial masks is one of the critical measures in combating the pandemic.

Figures 5 and 6, which demonstrate the effect of dynamic behavior by susceptible and symptomatically infected individuals respectively, show that preventing the symptomatic infectious from spreading the disease is as important as preventing the susceptible population from getting the infection. When the behavior of susceptible and symptomatically infected individuals was analyzed separately from each other, it turned out that the peak of the epidemic curve generated by symptomatic infections willing to self-isolate was lower than the peak of the epidemic curve generated by the susceptibles who are in support of the lock-down or closure measures. Thus, it is essential to prevent people from violating quarantine and social isolation rules especially as young people have been throwing “coronavirus parties” [49]. These parties are hosted either to defy social distancing rules or to get infected in hope to possibly build up immunity against the virus or simply because some people still think the virus is a hoax [39, 49].

One of our key findings is the possibility of multiple waves of infections due to rational human behavior. We saw in Figure 9 that these waves can persist when the rate of social learning of infected individuals is too high and their sensitivity to self-isolation is low. In this case, the infected individuals switch their behavior from self-isolating to not self-isolating while the prevalence of the infection is still relatively high; this results in a next wave of infections. The population quickly recognizes this shift in the state of the pandemic, and starts to self-isolate more often, thus suppressing this wave and repeating the cycle several times. On the other hand, the effect of such sensitive behavior can be mitigated by increasing quarantine and hospitalization rates (Figure 10).

Our key findings further show that when the symptomatic infectious population learn the positive behavior or are more willing to self-isolate, the community benefits, even though this change in behavior comes at a cost to them. Self-isolation often comes with financial implications and distress; not very many people can bear these burdens. Hence, it is important to incentivize self-isolation of the symptomatic infectious population as many infected people will rather stay home than go to work since staying at home will help the public good and create an opportunity to help save more lives [26]. One way to incentivize the symptomatic infectious is to pay them to stay home, perhaps via direct government subsidies for sick leave for infected individuals [26]. Our result shows that infection in the community will reduce particularly if the associated cost of self-isolation is cheap. If this cost is high and people keep violating quarantine rules, the infection could run away and become a persistent recurrent infection in the community, as shown in Figures 9–11.

We assumed that sensitivity to societal isolation measures was constant. However, public perception of these measures as necessary for the common good may change with time. For example, it may become a social norm to self-isolate in the face of a pandemic, and in this case infected individuals are more willing to isolate themselves from the rest of the population. A future iteration of this model should consider the effect of evolving public perception of the social stigma for those who refuse to self-isolate. We also considered the quarantine violation as a static feature of the model. However, the quarantine violation behavior may evolve with time just as self-isolating behavior. Constructing a dynamic game model of evolving quarantine violation behavior could involve an adaptive dynamic approach.

Additional concerns should be given to the ability to self-isolate. Proscriptive guidelines and current policies often fail to recognize that certain populations are less able or willing to stay at home due to compromised living situations, financial limitations, or precarious economic opportunities. Further approaches should consider how individual behaviors vary across key socioecomic and demographic population characteristics.

### 3.2 Conclusions

The goal of this study was to provide insight into possible effects of human behavior on non-pharmaceutical intervention strategies (such as partial lock-down and social isolation) aimed at containing the spread of COVID-19. Standard epidemiological models neglect human behavior, yet it is a major factor for studying COVID-19 transmission while there are no known pharmaceutical solutions. We showed that in certain circumstances rational human behavior may result in multiple waves of the pandemic, which persist for a long period of time.

Finally. we summarize our results according to whether human behavior is static or dynamic driven by public perception of risk of the infection and sensitivity to isolation measures.

a. The simulations of the COVID-19 model (1) with static human behavior (constant quarantine violation rate) show that:
  i. Increased quarantine violation and discharge rates of those still infectious due to overwhelmed hospital resources results in greater disease burden leading to an early epidemic peak.
  ii. Increasing quarantine and hospitalization rates reduces the disease burden and the epidemic peak.
b. The simulations of the COVID-19 model (15) with dynamic human behavior show that:
  i. Symptomatic individuals learning and mimicking positive behavior reduces the disease burden in the population but can lead to multiple epidemic waves if fewer susceptible individuals mimic and learn positive behavior.
  ii. Quarantine violation and hospital discharge of symptomatic individuals amplifies the peaks of the infection waves and can lead to infection waves that persist in the community.
  iii. Increasing quarantine and hospitalization rates can prevent multiple waves of infection.
  iv. It is important to incentivize the burden of self-isolation to encourage more symptomatic infectious to self-isolate because high cost of self-isolation is not beneficial to the infectious nor to the community as a whole.

Overall, our results emphasize the importance of diverse steps that could be implemented that would incentivize and support responsible behavior by individuals. This might involve positive reinforcement, such as subsidies and economic support, or negative consequences, such as penalties and fines for those not obeying and following appropriate behavioral norms.

## 4 Methods

In this study, we develop a novel COVID-19 transmission model that incorporates dynamic human behavior, which is driven by various factors. We parameterized the model using data from the ongoing COVID-19 outbreaks. To develop this novel game-theoretic model with dynamic human behavior, we first consider a baseline epidemiological model with static human behavior.

### 4.1 Baseline COVID-19 model

We construct a model of COVID-19 transmission with quarantine and hospitalization. We follow the natural history of the infection [42, 53] and partition the population according to their disease status as susceptible (*S*(*t*)), exposed (*E*(*t*)), asymptomatically infected (*A*(*t*)), symptomatically infected (*I*(*t*)),quarantined (*Q*(*t*)), hospitalized (*H*(*t*)), and removed (*R*(*t*)) individuals. The static human behavior in this model is represented by the constant rate of violating quarantine.

We assume that the population is not affected by birth and natural mortality because we are modeling short-term dynamics of the pandemic. We therefore treat compartment sizes as proportions of the entire population. Susceptible individuals become exposed upon contact with infected individuals, and the force of infection is given by

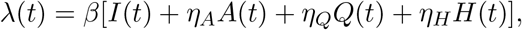

where *β* is the infection rate, and *η_A_*, *η_Q_*, and *η_H_* are the modification parameters representing reduced infectiousness of asymptomatic, quarantined, and hospitalized individuals respectively.

Exposed individuals become infected at the rate *σ*. A proportion *q* of these individuals show no symptoms of the disease and move to the asymptomatically infected compartment, while a proportion (1−*q*) of exposed individuals develop clinical symptoms of the disease and move to the symptomatically infected compartment. Asymptomatic (symptomatic) individuals recover from the disease at the rate *γ_A_* (*γ_I_*) and die at the rate *δ_A_* (*δ_I_*). Symptomatic individuals are hospitalized at the rate *ω_H_*. Those individuals whose condition is not sufficiently severe are quarantined at the rate *ω_Q_*. There have been reports of people flouting quarantine [15,19,25,38], and we assume that quarantined individuals break the quarantine at the rate *ν_Q_*. Quarantined individuals recover from the disease at the rate *γ_Q_* and die at the rate *δ_Q_*.

COVID-19 spreads at an alarming rate, requiring high rates of hospitalization. Hospitals often become overwhelmed and may run out of beds, respirators, ventilators, and ICUs [47]. Furthermore, some hospitals are reserving beds for the critically ill COVID19 patients and discharging those with less severe illness [1,27]. We assume that due to the limitations in hospital capacity, hospitalized individuals leave the hospitals while still infected at the rate *ν_H_*. Hospitalized individuals recover from the disease at the rate *γ_H_* and die at the rate *δ_H_*.

The removed individuals comprise both recovered and deceased individuals. We disregard the possibility of reinfection because we are looking into short-term dynamics of the disease spread in the population. We therefore assume that recovered individuals do not contribute to the spread of the infection.

The flow diagram depicting the transitions between compartments as the disease progresses through the population is shown in Figure 1, and the associated state variables and parameters are described in Table 1.

The differential equations describing the dynamics of this model are given in Equation (1).

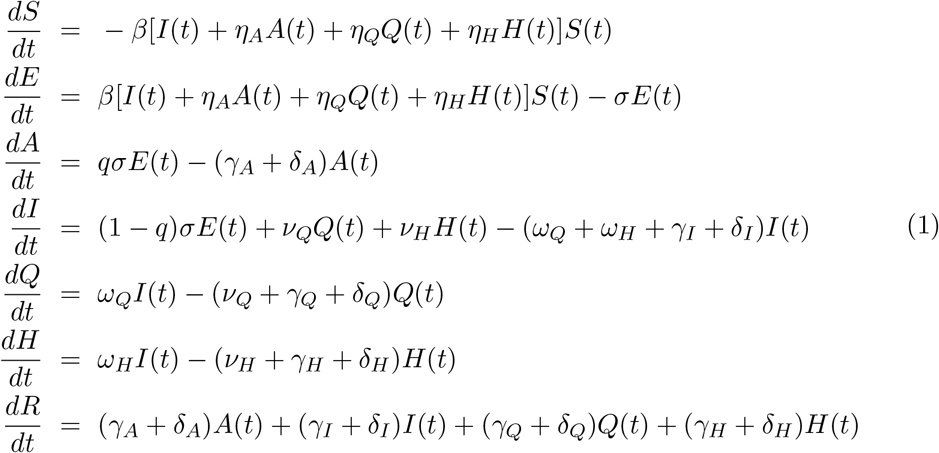

### 4.2 Model of dynamic human behavior

In this section, we use the imitation dynamic approach of evolutionary game theory [3, 43] to model evolving human behavior in response to the pandemic and its effect on the spread of the disease. We consider behavioral response of both susceptible and infected individuals. Susceptible individuals wish to protect themselves from getting infected, and they consider supporting social distancing measures such as school and workplace closures. On the other hand, conscientious infected individuals consider self-isolation as means to protect the rest of the population. We begin by modeling each type of behavior separately, and then we implement both behavioral responses within our baseline COVID-19 model.

#### 4.2.1 Susceptible individual support for school and workplace closure

As the pandemic rages on without any known pharmaceutical drugs or vaccines, using personal protection equipment (PPE), washing hands, social distancing, and economic lock-downs are the measures recommended to contain and control the disease [12,30, 52]. We adopt the approach of [41] to model the behavioral response of the susceptible individuals. The susceptible individuals have two strategies to choose from: to support closure or not to support closure; we let *x_S_* (*t*) denote the proportion of susceptible individuals that support closure. The time-varying function *C*(*t*)captures the impact of social distancing measures such as school and workplace closure on the transmission of COVID-19. The evolution of the susceptible and exposed sub-populations with social distancing becomes

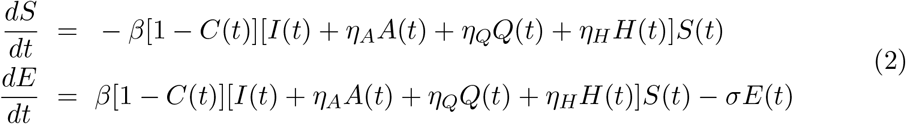

Following [41], we define

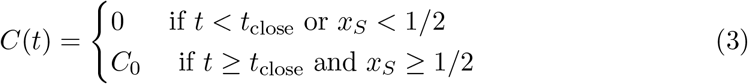

where *C*_0_ is a combined measure of the effectiveness of physical distancing in those workplaces that remain open and how many workplaces are closed. The decision to close schools and workplaces is “turned on” if the time after the start of the pandemic is at least *t*_close_ and at least half of the (susceptible) population supports closure. The closure policy is “turned off” if less than half of the (susceptible) population supports closure.

The susceptible individuals weigh the risk of the infection based on the disease prevalence and the accumulating socio-economic losses due to the closures. The susceptible individuals who do not support school and workplace closure are willing to face the risk of infection, and their perceived payoff is given by

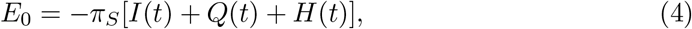

where *π_S_* is the sensitivity to being infected with COVID-19 parameter. The susceptible individuals who support closure efforts face socio-economic losses, and their perceived payoff is given by

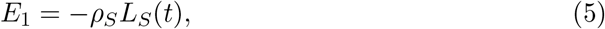

where *ρ_S_* is the the sensitivity to the accumulated socio-economic losses *L_S_* (*t*), as in [41].

We now describe how the behavioral responses of susceptible individuals evolve with time. An individual who did not support closure but decided to switch its strategy achieves a payoff gain

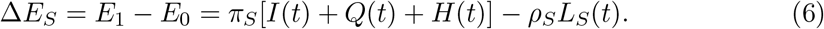

We assume that individuals employ a social learning process where they adopt strategies of other individuals with the rate proportional to the payoff gain, which can be realized via an imitation dynamic. The proportion of susceptible individuals who support closure thus evolves according to

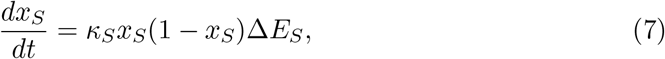

where *κ_S_* is the social learning rate. The individuals who do not support closure (1−*x_S_*) sample the individuals who do support closure (*x_S_*) and switch their strategy at the rate proportional to the payoff gain *ΔE_S_*. Using equation (6), we obtain

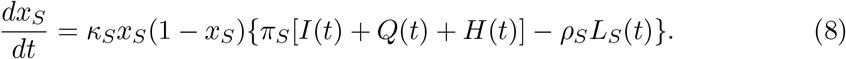

Individuals are thus more likely to support closure if the prevalence of the infection is high and/or socio-economic losses due to the closures are low. On the other hand, due to the accumulating nature of the socio-economic losses, individuals are not likely to support closure for too long.

Since scaling payoff functions does not affect the outcome, we can replace Δ*E_S_* given by (6) with Δ*E_S_* = *I*(*t*)+*Q*(*t*)+*H*(*t*)−(*ρ_S_ /π_S_*)*L_S_* (*t*). Then

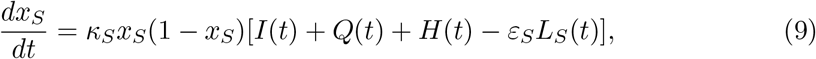

where *ε_S_* = *ρ_S_ /π_S_* is the sensitivity to the socio-economic losses relative to getting infected with COVID-19.

Finally, following [41], the evolution of the time-varying quantity *L_S_* (*t*), which represents the accumulated socio-economic losses, obeys the exponential fading memory mechanism given by

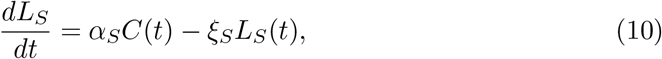

where *α_S_* controls the rate at which school and workplace closures impacts socioeconomic health of the population, and *ξ_S_* is a decay rate that represents adjustment to the baseline losses.

#### 4.2.2 Infected individual self-isolation

While susceptible individuals seek to avoid getting infected, the symptomatically infected individuals cannot help themselves. We thus assume that conscientious symptomatically infected individuals seek to minimize the potential damage to the susceptible part of the population.

Since COVID-19 was elevated to pandemic status, self-isolation and quarantine had been the prescribed non-pharmaceutical measures aimed at flattening the incidence curve. China (at the peak of infection) instituted mandatory quarantine of individuals and some parts of the country[24,32]. Other countries imposed travel bans and recommended 14-day quarantines (via self-isolation) for their citizens who travel to hotspot places [40, 50, 51]. However, people break and violate self-isolation and quarantine [19, 38]either due to quarantine fatigue or to other factors such as procuring material needs or limited opportunities to maintain isolation [7,17]. Some have engaged in even more deadly behaviors ignoring policies and attending large social gatherings [21,49].

We assume that the symptomatically infected individuals who tested positive for COVID-19 and were ordered to quarantine themselves leave quarantine at a constant rate *ν_Q_*. However, symptomatically infected individuals (*I*(*t*)) whose condition was not severe enough to go to a hospital and/or get tested may elect to self-isolate to protect others. Let *x_I_* (*t*) be the proportion of symptomatically infected individuals *I*(*t*) who elect to self-isolate. We assume that self-isolated individuals do not contribute to the spread of the infection, and the force of infection term involving *I*(*t*) becomes (1−*x_I_* (*t*))*I*(*t*). Hence, the equations for the susceptible and exposed individuals from the baseline model become

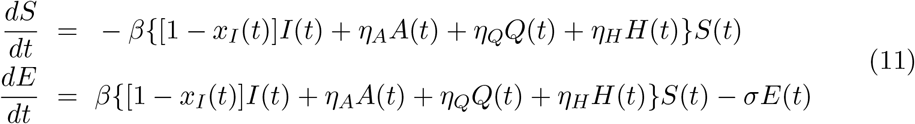

A symptomatically infected individual who elects not to self-isolate faces the burden of infecting other individuals. These individuals use the publicly available information on the COVID-19–induced death rates to estimate the extent of the burden. We therefore assume that the payoff of an individual who chooses not to self-isolate is given by

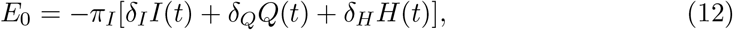

where *π_I_* is the sensitivity to infecting others parameter. On the other hand, an infected individual who decides to self-isolate faces a fixed cost of such a decision because the length of self-isolation is approximately equal to the time it takes to recover. Hence, the payoff of an individual who chooses to self-isolate is given by

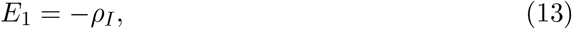

where *ρ_I_* is the sensitivity to self-isolation parameter.

Similar to the closure support model described above, the proportion of symptomatically infected individuals who elect to self-isolate evolves according to the imitation dynamic

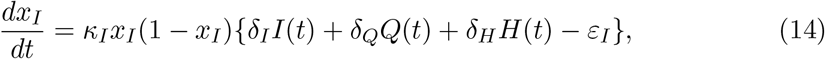

where *κ_I_* is the self-isolation social learning rate, and *ε_I_* = *ρ_I_ /π_I_* is the sensitivity to self-isolation relative to infecting others. The (conscientious) infected individuals would tend to self-isolate if theCOVID-19–induced death toll is high, while they would tend not to self-isolate as long as the death rates become sufficiently low.

#### 4.2.3 The COVID-19 model with combined dynamic behavior

We now combine the two types of adaptive strategic responses in the population. The susceptible individuals elect to either support or not support school and workplace closures, while infected individuals elect to self-isolate or not to self-isolate. Combining equations (2) and (11) and replacing the corresponding equations in the base line model (1) results in a coupled COVID-19 model with combined behavioral effects where parts of the population adjust their behavior after sampling or learning other people’s behavior according to the appropriately defined payoffs. This coupled disease-behavior system is given by the following system of ordinary differential equations:

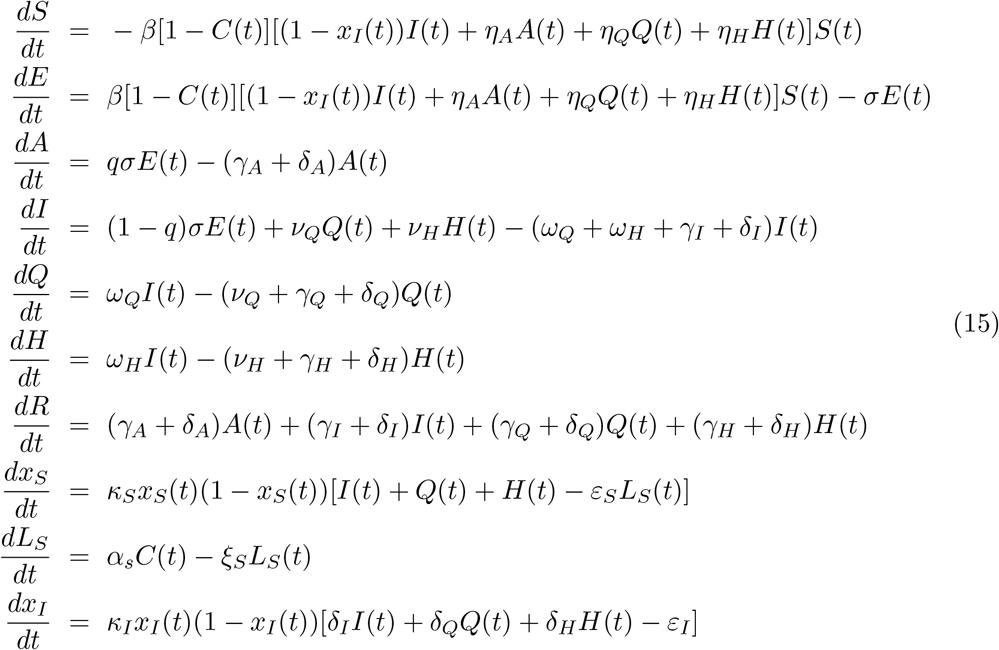

The game-theoretic model of dynamic human behavior state variables and parameters are summarized in Table 2.

**Table 2:** The dynamic human behavior model state variables and parameters.

| Variable | Description |
| --- | --- |
| $x_S(t)$ | Proportion of susceptible individuals who support closure |
| $x_I(t)$ | Proportion of symptomatically infected individuals who self-isolate |
| $C(t)$ | Impact of school and workplace closures |
| $L_S(t)$ | Accumulated socio-economic losses due to closures |
| Parameter | Description |
| $\kappa_S$ | Support for closure social learning rate |
| $\kappa_I$ | Self-isolation social learning rate |
| $\varepsilon_S$ | Sensitivity to socio-economic losses relative to COVID-19 infection |
| $\varepsilon_I$ | Sensitivity to self-isolation relative to infecting others |
| $t_{\text{close}}$ | Initial time closures may take effect |
| $C_0$ | Effectiveness of the closure measures |
| $\alpha_S$ | Closure impact rate on socio-economic health |
| $\xi_S$ | Decay rate for socio-economic losses |

### 4.3 Data and model fitting

We obtained COVID-19 cumulative number of cases data for Arizona, for a period of time from January 26 to July 6, 2020, from the Johns Hopkins website [31] and fitted it to thebaselineCOVID-19 model(1) to estimate the values of the model parameters; see Figure 12.

**Figure 12:**
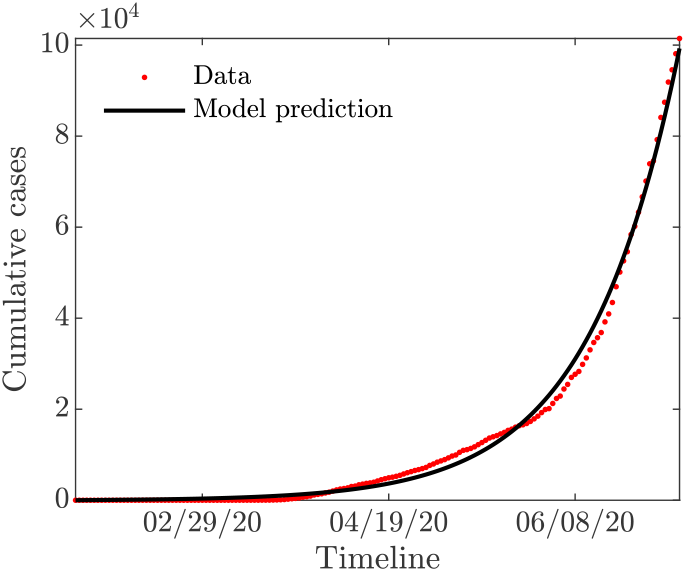
Fitting the baseline COVID-19 model parameters (1) to Arizona data of reported cumulative new cases. The COVID-19 outbreaks data are obtained from Johns Hopkins website [31].

The values of the baseline model parameters are summarized in Table 3. We used these values to estimate the value of 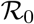 for the COVID-19 outbreak in Arizona as 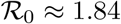.

**Table 3:** Parameters values for the baseline COVID-19 model (1) fitted to Arizona.

| Parameter | Description | Value | References |
| --- | --- | --- | --- |
| $\beta$ | Infection rate | 0.4712 | Fitted |
| $\eta_A$ | Asymptomatic infection rate modification parameter | 0.45 | [9] |
| $\eta_Q$ | Quarantined infection rate modification parameter | 0.0101 | Fitted |
| $\eta_H$ | Hospitalized infection rate modification parameter | 0.4509 | Fitted |
| $q$ | Proportion developing asymptomatic infections | 0.5 | [9] |
| $\sigma$ | Disease progression rate | 1/6 | [9] |
| $\gamma_I$ | Recovery rates of symptomatic | 0.5997 | Fitted |
| $\gamma_A$ | Recovery rates of asymptomatic | 0.2363 | Fitted |
| $\gamma_Q$ | Recovery rates of quarantined | 0.3815 | Fitted |
| $\gamma_H$ | Recovery rates of hospitalized | 0.0107 | Fitted |
| $\omega_Q$ | Quarantine rate | 0.5326 | Fitted |
| $\omega_H$ | Hospitalization rate | 0.7495 | Fitted |
| $\nu_Q$ | Quarantine violation rate | 0.4586 | Fitted |
| $\nu_H$ | Hospital discharge rate | 0.0126 | [9] |
| $\delta_I$ | Death rate of symptomatic | 0.0065 | Fitted |
| $\delta_A$ | Death rate of asymptomatic | 0.00325 | Assumed |
| $\delta_Q$ | Death rate of quarantined | 0.0065 | [9] |
| $\delta_H$ | Death rate of hospitalized | 0.0065 | [9] |

## Data Availability

Data used are included in the text.

## Acknowledgements

This research is supported by the National Science Foundation under grant number DMS 2028297. FBA would like to thank Chris Bauch for sharing their second wave imitation dynamics code.

## A Appendix A: Contour plots of theCOVID-19 reproduction number 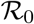

**Figure 13:**
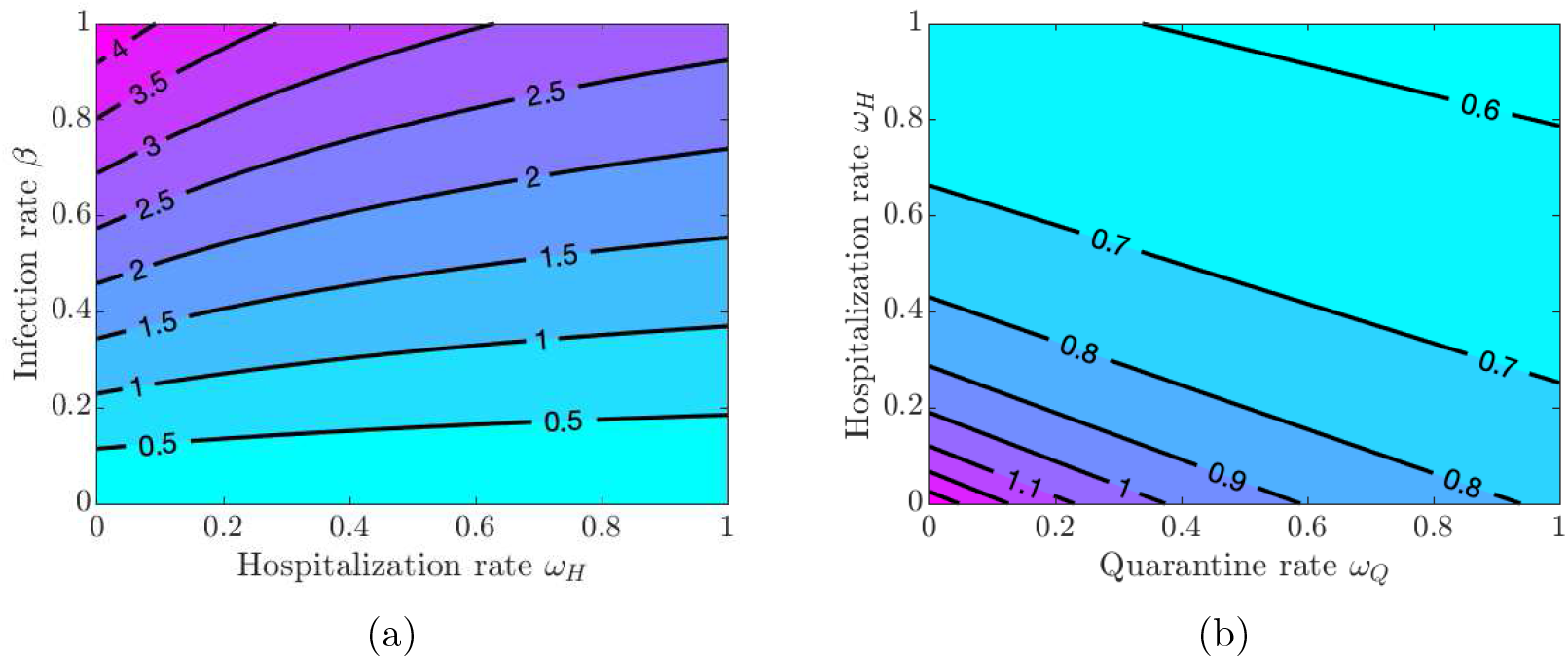
Contour plot of the COVID-19 reproduction number 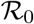 given in equation (1). (a) Varying hospitalization rate *ω_H_* and infection rate *β*. (b) Varying quarantine rate *ω_Q_* and hospitalization rate *ω_H_* using infection rate *β* =0.22.

**Figure 14:**
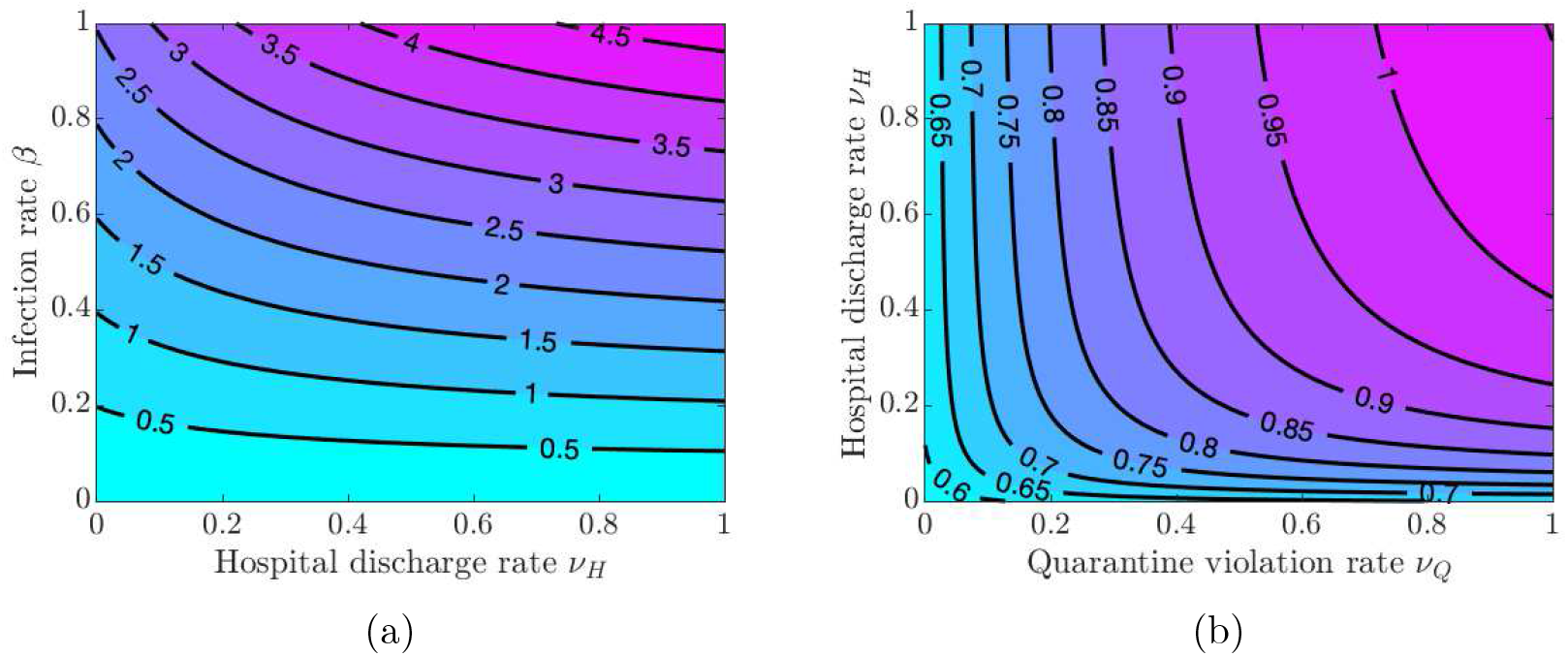
Contour plot of the COVID-19 reproduction number 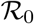 given in equation (1). (a) Varying quarantine violation rate *ν_Q_* and infection rate *β*. (b) Varying quarantine violation rate *ν_Q_* and hospital discharge rate *ν_H_* using infection rate *β* =0.22.

## B Appendix B: Impact of high cost self-isolation (*ε_I_*) on symptomatic infectious

Here we show the simulation results for all symptomatic infections with high sensitivity to self-isolation *ε_I_* =0.00008.

**Figure 15:**
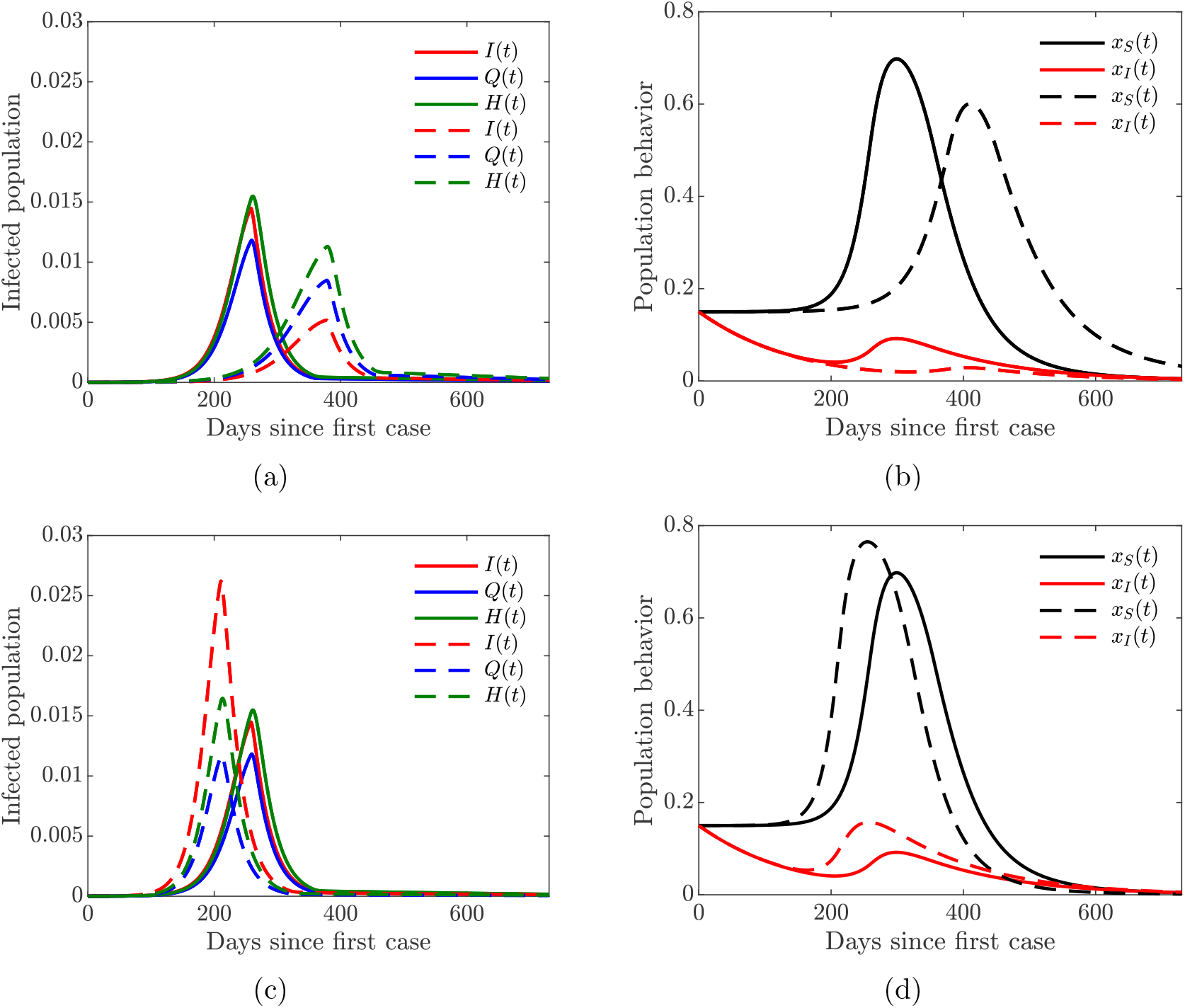
Simulations of theCOVID-19 model with dynamic human behavior(15) for the proportions of all symptomatic infections and behavioral response with high sensitivity to self-isolation *ε_I_* =0.00008. The social learning rates are *κ_S_* = 1 and *κ_I_* =100, and *x_S_* (0)= *x_I_* (0)=0.15. Solid lines correspond to the values of the baseline model parameters given in Table 3. (a)–(b)Dashed lines correspond to double quarantine (*ω_Q_*) and hospitalization (*ω_H_*) rates (c)–(d) Dashed lines correspond to double quarantine violation (*ν_Q_*) and hospital discharge (*ν_H_*) rates.

## C Appendix C: The impact of symptomatic social learning rates *κ_I_*

Here we show the simulation results of varying symptomatically infected individuals social learning rate *κ_I_* with high sensitivity to self-isolation *ε_I_* =0.00008.

**Figure 16:**
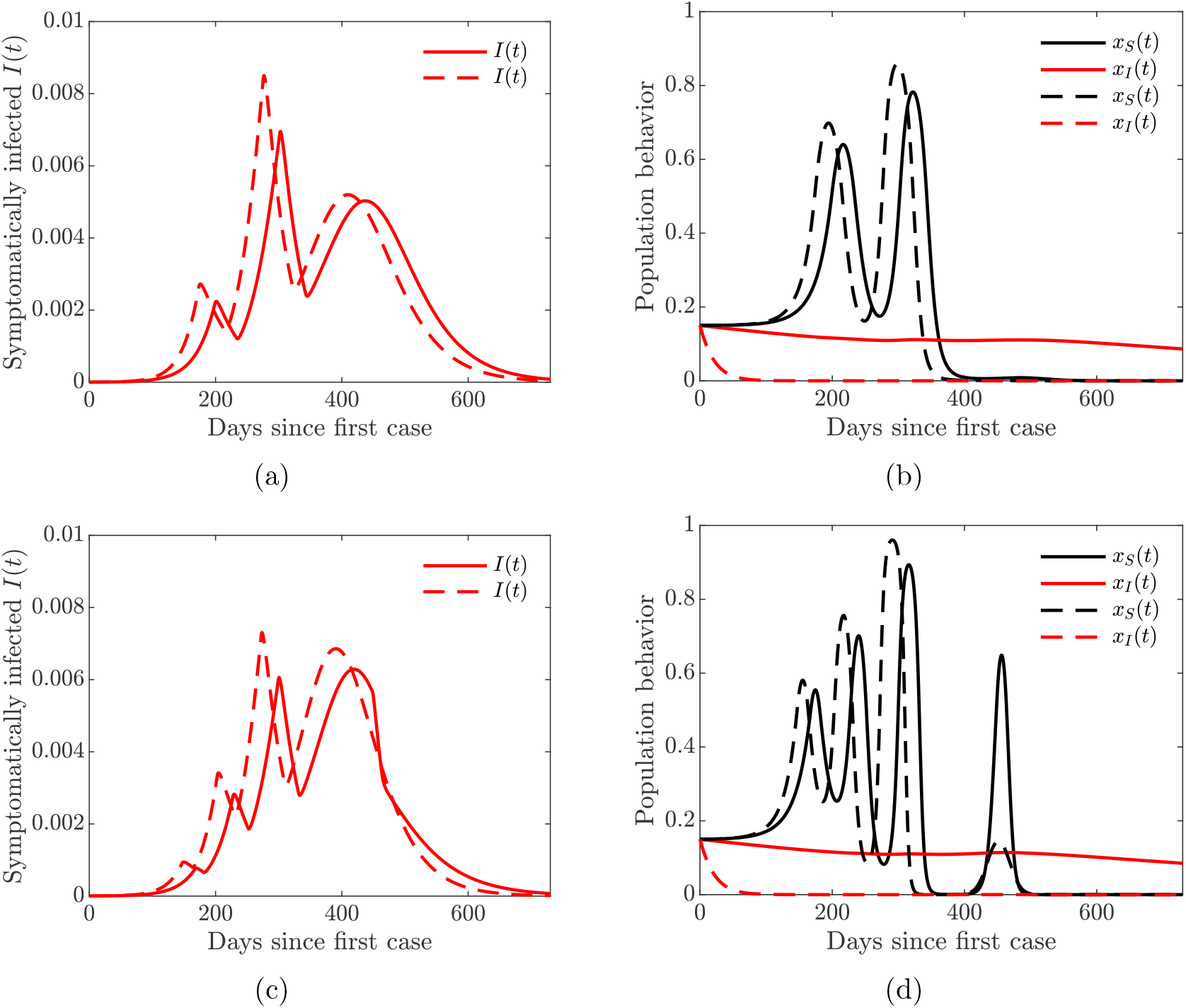
Simulations of the COVID-19 model with dynamic human behavior (15) for the proportions of all symptomatic infections and behavioral response with high sensitivity to self-isolation *ε_I_* =0.00008. Solid lines correspond to *κ_I_* = 20, dashed lines correspond to *κ_I_* = 650. (a)–(b) *κ_S_* =10;(c)–(d) *κ_S_* =30.

